# T cell responses in repeated controlled human schistosome infection compared to natural exposure

**DOI:** 10.1101/2025.02.13.25322130

**Authors:** Emmanuella Driciru, Jan Pieter R. Koopman, Sanne Steenbergen, Friederike Sonnet, Koen A. Stam, Helena de Bes-Roeleveld, Eva Iliopoulou, Jacqueline J. Janse, Jeroen Sijtsma, Irene Nambuya, Stan T. Hilt, Marion König, Yvonne Kruize, Miriam Casacuberta-Partal, Moses Egesa, Govert J. van Dam, Paul L.A.M. Corstjens, Lisette van Lieshout, Harriet Mpairwe, Andrew S. MacDonald, Maria Yazdanbakhsh, Alison M. Elliott, Meta Roestenberg, Emma L. Houlder

## Abstract

Schistosomiasis affects over 250 million people, predominantly in Sub-Saharan Africa. In *Schistosoma*-endemic regions a lack of natural sterilizing immunity means individuals are repeatedly infected, treated and reinfected. Due to difficulties in tracking infection in endemic areas, the kinetics of the host immune response during these reinfections have not been elucidated. Here, we use repeated (3x) controlled human *Schistosoma mansoni* (*Sm*) infection (repeated CHI) to directly study how antigen-specific T cells develop during reinfection. We compared these responses to natural *Sm*-infected endemic Ugandan individuals. We report that a mixed Th1/Th2/regulatory CD4^+^ T cell response develops in repeated CHI, primarily against adult worm antigens. Adult worm-specific responses after repeated CHI were similar to those observed in naturally infected endemic participants. However, endemic participants showed differential responses to antigens from the egg and cercariae life-cycle stages. Repeated CHI (3x) with sequential exposure to *Sm* cercariae of different sexes (male-female-male) revealed an elevated CD4^+^ T cell cytokine response to adult worm and egg antigens. Our findings demonstrate that reinfection with single-sex schistosomes elicits adult worm-specific T cell cytokine responses that reflect endemic-natural infection, highlighting the translatability of the CHI model to natural infection in endemic settings. Overall, this study advances our understanding of how schistosome immune responses develop over reinfection cycles in the human host, thereby increasing our understanding of the immunology of natural schistosome (re)infection.

## Introduction

Schistosomiasis is a neglected tropical disease caused by infection with helminths of the *Schistosoma* species. Over 250 million people are affected globally, with the Sub-Saharan region accounting for more than 90% of infections (*1*). Continuous infection with the long-lived *Schistosoma* parasites exerts a chronic immune modulatory effect on the host due to prolonged exposure to schistosome antigens, primarily via prolific egg production (*2*). Existing evidence on host immune response is mainly derived from murine studies, and shows a distinct immune trajectory across infection stages, with a T helper 1 (Th1) phenotype followed by Th2 and regulatory responses post egg production (*3–5*). Egg-induced host immune responses are responsible for disease pathology and morbidity, although only 10% of cases progress to severe disease (*6*).

Reinfection due to repeated exposure, usually from early childhood, is common in endemic settings. Although praziquantel (PZQ) chemotherapy effectively reduces worm burden to undetectable levels, reinfection rapidly occurs. A recent study showed prevalence is restored to over 44.5% within 8 months of PZQ administration among shoreline communities of Lake Victoria, Uganda (*7, 8*). Over time, individuals in the highly endemic areas show reduced infection burden with age, after repeated infection-treatment cycles, attributable to partial immunity and/or altered exposure patterns (*9, 10*). Increased Th2 cytokines, high eosinophil count, and worm-specific IgE levels, have been associated with natural immunity, although not all are consistently found in meta-analyses (*4, 9, 11–13*). How the immune response develops during natural repeated infection-treatment cycles, potentially leading to partial immunity, is still unclear. Natural infection studies are inherently limited by asynchronous and diverse exposure histories, hindering understanding of immune kinetics, which requires a precise definition of (re-) infection time points, dose, interval, and duration.

With the advent of controlled human challenge infection (CHI) studies, it is now possible to probe in depth into infection immune dynamics using a precise longitudinal approach. In our previous single-sex single-infection *Schistosoma*-CHI model, Dutch participants showed symptom-associated inflammatory Th1 responses at 4 weeks post-infection, followed by Th2/regulatory responses at week 8 (*14*). Notably, the Th2 response developed despite the absence of eggs, previously considered to be crucial for the switch to a Th2 response (*15, 16*).

To increase translatability of the CHI model to the endemic situation we designed a repeated *Schistosoma mansoni* (*Sm*) CHI model, exposing *Sm*-naïve Dutch volunteers to three cycles of cercariae and PZQ treatment (*17*). Reinfection did not induce immunity, but symptoms decreased after the second and third exposure in the reinfection group (*17*). During this trial, routine stool PCR testing and confirmatory microscopy after the third exposure unexpectedly detected a low number of *Sm* eggs in one participant. No eggs were detected in any other participants. A retrospective examination found that during the second exposure, five of twelve volunteers were unintentionally exposed to female cercariae (*17*). These five volunteers received male-female-male (m-f-m) exposures at the first, second, and third time points, respectively, in contrast to the planned male-male-male (m-m-m) exposure (Fig. 1A). The possibility of egg production after the third exposure in the m-f-m exposed participants could have resulted from ineffective clearance of female worms by PZQ, allowing for worm pairing after the third (male) exposure (*18*). During the third exposure, m-f-m exposed volunteers (with potential eggs) tended to have higher eosinophil count, antibody and serum cytokine levels (*17*).

**Figure 1:**
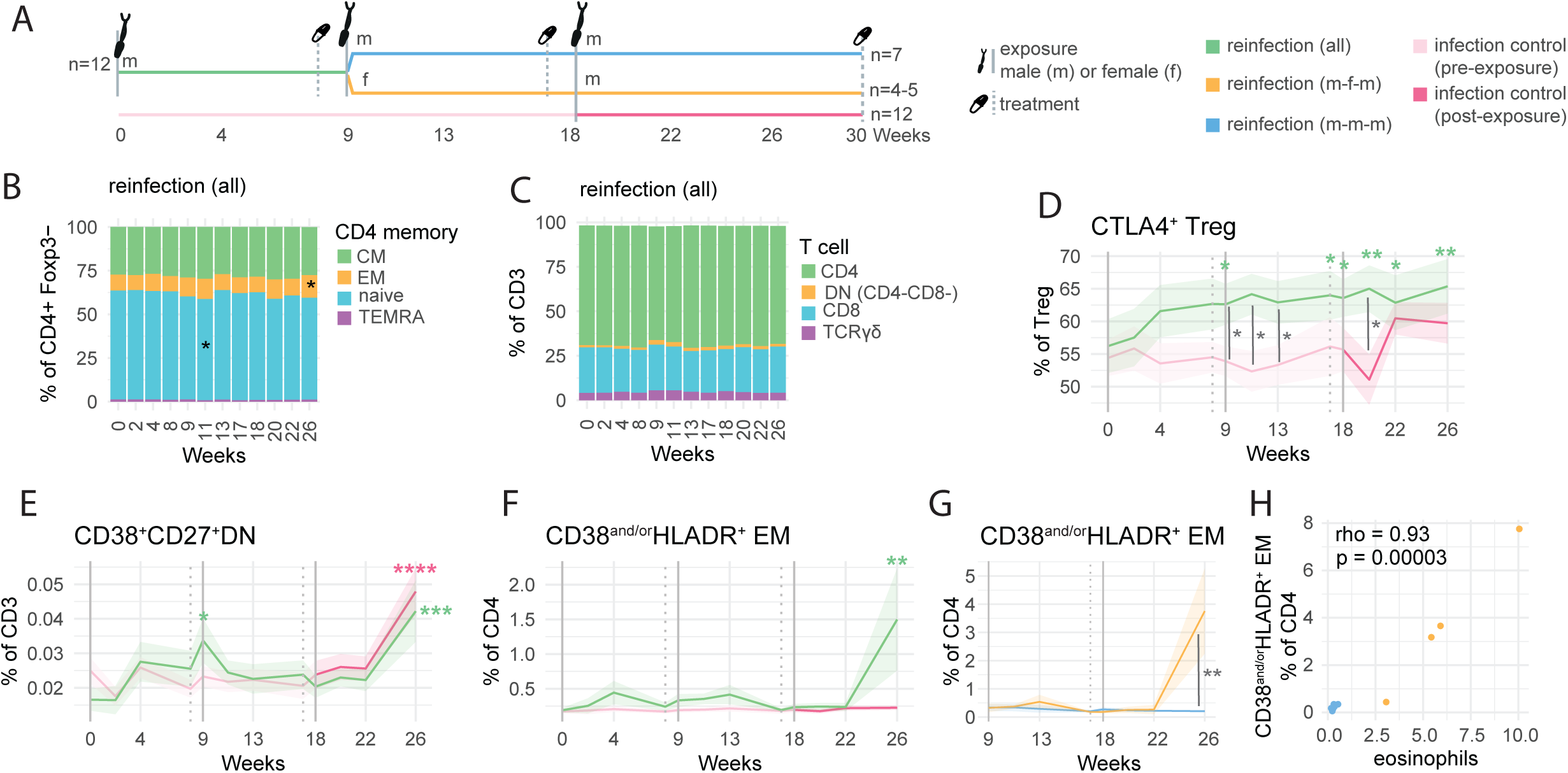
Subtle alterations to T cell phenotype during repeat schistosome infection. A) Overview of trial design. Volunteers were randomised to reinfection (n=12) or infection control (n=12) groups. Reinfection volunteers (green) underwent 3 exposure (weeks 0,9,18) and treatment (weeks 8,17,30) cycles, whereas the infection control group (pink) underwent a single exposure (at week 18). At the second infection n=5 volunteers were unintentionally exposed to female cercariae (f) with groups referred to from then onwards as reinfection male-female-male (m-f-m, yellow) or reinfection male-male-male (m-m-m, blue). B) Stacked bar chart of CD4 T cell memory frequencies. Each bar section represents the mean frequency. C) Stacked bar chart of T cell subset frequencies. Each bar section represents the mean frequency. D-F) Ribbon plots depicting mean (lines) frequency and standard error (shaded area) of the mean of specified populations, coloured by infection group. In panels B-F a linear mixed model was performed to (separately) assess changes in the reinfection and infection control group compared to the week 0 baseline, with volunteer as a random effect. FDR values are displayed in black in figure C, and green (reinfection) and pink (infection control) in figures D-F. * FDR <0.05, ** FDR<0.01, ***FDR<0.001, ****FDR<0.0001. To compare between reinfection (all) and infection control groups welch’s T-tests were performed at each timepoint with significant p values (uncorrected) displayed with a grey line * p <0.05. G) Ribbon plots depicting mean (lines) frequency and standard error (shaded area) of the mean of CD38and/orHLADR+ EM CD4 T cells in the reinfection (m-m-m) and reinfection (m-f-m) groups. To compare between reinfection (m-m-m) and reinfection (m-f-m) groups Mann-Whitney tests were performed at each timepoint with significant p values (uncorrected) displayed with a grey line * p <0.05, ** p <0.01. H) Scatter plots comparing CD38and/orHLADR+ EM CD4 T cell frequency to blood eosinophilia at week 26. Spearman’s rank correlation analysis was performed with the correlation coefficient rho and p value reported.

Utilizing samples from this repeated CHI model, we report how immune responses develop during repeated *Sm* infection and treatment cycles, a study designed to mimic endemic exposure patterns. By directly comparing samples from repeated CHI in Dutch volunteers to naturally infected Ugandan participants we evaluate how translatable findings in the CHI model are to natural endemic infection. Using spectral flow cytometry to analyze cellular responses in peripheral blood mononuclear cell (PBMC) samples, we observed a mixed Th1/Th2/regulatory response developed in repeated CHI, primarily directed against adult worm antigens (AWA). The T cell response in repeated CHI is comparable in magnitude to that in endemic schistosome infection but is reduced when compared to repeated CHI volunteers that were sequentially exposed to schistosome cercariae of different sexes (m-f-m exposure).

## Results

### Single-sex repeated infection challenge alters the CD4+ T cell phenotype

To investigate cellular changes in the T cell compartment induced by repeated *S. mansoni* CHI (Fig. 1A), PBMC samples were analyzed using a 30+ marker spectral flow cytometry panel. At week 11 (two weeks post the second exposure), there was a significant reduction in the frequency of naïve CD4 + T cells in the reinfection (all) group compared to baseline (week 0) (Fig. 1B). Such decrease may indicate possible onset of adaptive responses following PZQ-induced worm antigen unmasking (*19–21*). Moreover, at week 26, effector memory (EM) T cells increased in the reinfection (all) group (Fig. 1B). Subsets in the total CD4+ T cell compartment – CD4^+^ T cells, CD8^+^ T cells, γδT cells or DN (double negative CD4^-^CD8^-^) T cells – remained unchanged (Fig. 1C). In the infection control group that received a single exposure at week 18, no significant alterations in any T cell populations were observed (Supp. Fig. 1A-B)

Next, we examined changes in the CD4 T cell activation status. By week 9, CTLA4^+^ regulatory T cells increased significantly in the reinfection (all) group compared to week 0 baseline, staying elevated throughout the second and third infections (Fig. 1D). Moreso, at weeks 9, 11, 13, and 22 this regulatory T cell subset was significantly higher in the infection group compared to infection control group which did not show any significant increases (Fig. 1D). Similarly, at week 9 and 26 a CD38^+^CD27^+^ DN T cells increased significantly in the reinfection (all) group, as well as week 26 in the infection control group. This population is thought to be regulatory, and has been previously shown to increase after single *Sm* CHI (*14*).

At week 26 (8 weeks post third exposure) activated CD38^AND/OR^HLADR^+^ EM T cells significantly expanded in the reinfection (all) group (Fig. 1F) (*14*). Other CD4 T cell populations – CD4^+^ Treg, PD1^+^ Treg, CD38/HLADR^+^ Treg, Gata3^+^CRTH2^+^ CD4^+^ T cell, CTLA4^+^CD4^+^ Teg and PD1^+^CD4^+^ T cells - remained generally stable (Supp. Fig. 1C-H). When m-f-m and m-m-m reinfection groups were split, we observed that the expansion of CD38^AND/OR^HLADR^+^ EM T cells at week 26 was attributable to m-f-m volunteers, with potential egg production (Fig. 1G). Significant differences between the m-f-m and m-m-m groups were not observed in other CD4 T cell populations (Supp. Fig. 1I-P). Notably, CD38^AND/OR^HLADR^+^ EM T cell expansion significantly correlated with peripheral eosinophilia (*17*), suggestive of a concerted inflammatory/type-2 response in these m-f-m volunteers (Fig. H).

Overall, these results demonstrate modest increases in the regulatory cell (CTLA4^+^ Treg and DN T cell) populations in single-sex (m-m-m) reinfection. In m-f-m volunteers a striking increase in activated CD38^AND/OR^HLADR^+^ EM T cells was observed at week 26, following potential egg production.

### Repeated infection induces a mixed CD4^+^ T cell-specific cytokine response to adult worm antigen, but not cercarial antigen

To understand if repeated infection changes T cell cytokine response to the different schistosome life cycle stages, PBMCs were stimulated with adult worm (AWA) and cercarial (CA) antigens. A robust CD4^+^ T cell cytokine response was observed upon AWA stimulation in the reinfection (all) group. This response comprised of significant elevation in Th1, Th2, and regulatory cytokines - TNF, IFN-y, IL-13, IL-4, IL-9, IL-5, IL-10, IL-22, and IL-21 - compared to the baseline (Fig. 2A-G, Supp. Fig. 2A-E). AWA-specific TNF and IL-22 were significantly increased in the reinfection group over infection control during the first infection (wk 4-8) (Fig. 2F, Supp. Fig. 2C). Generally however, AWA-specific responses were most visible at week 11, after the first PZQ treatment was administered (Fig. 2A-G). This could be attributed to an immune boosting by the second exposure (2 weeks prior) and or antigen release due to PZQ treatment (3 weeks prior) (*22*).In contrast, a muted response to CA was observed in the reinfection (all) group, with significant increases occurring only in the IFN-y, TNF, IL-5, and IL-13 cytokine levels (Fig. 2H-J, Supp. Fig. 2F-N). As expected, changes in antigen-specific cytokine response in the infection control group were observed only after cercariae exposure at the third exposure timepoint (Supp. Fig. 2A&F).

**Figure 2:**
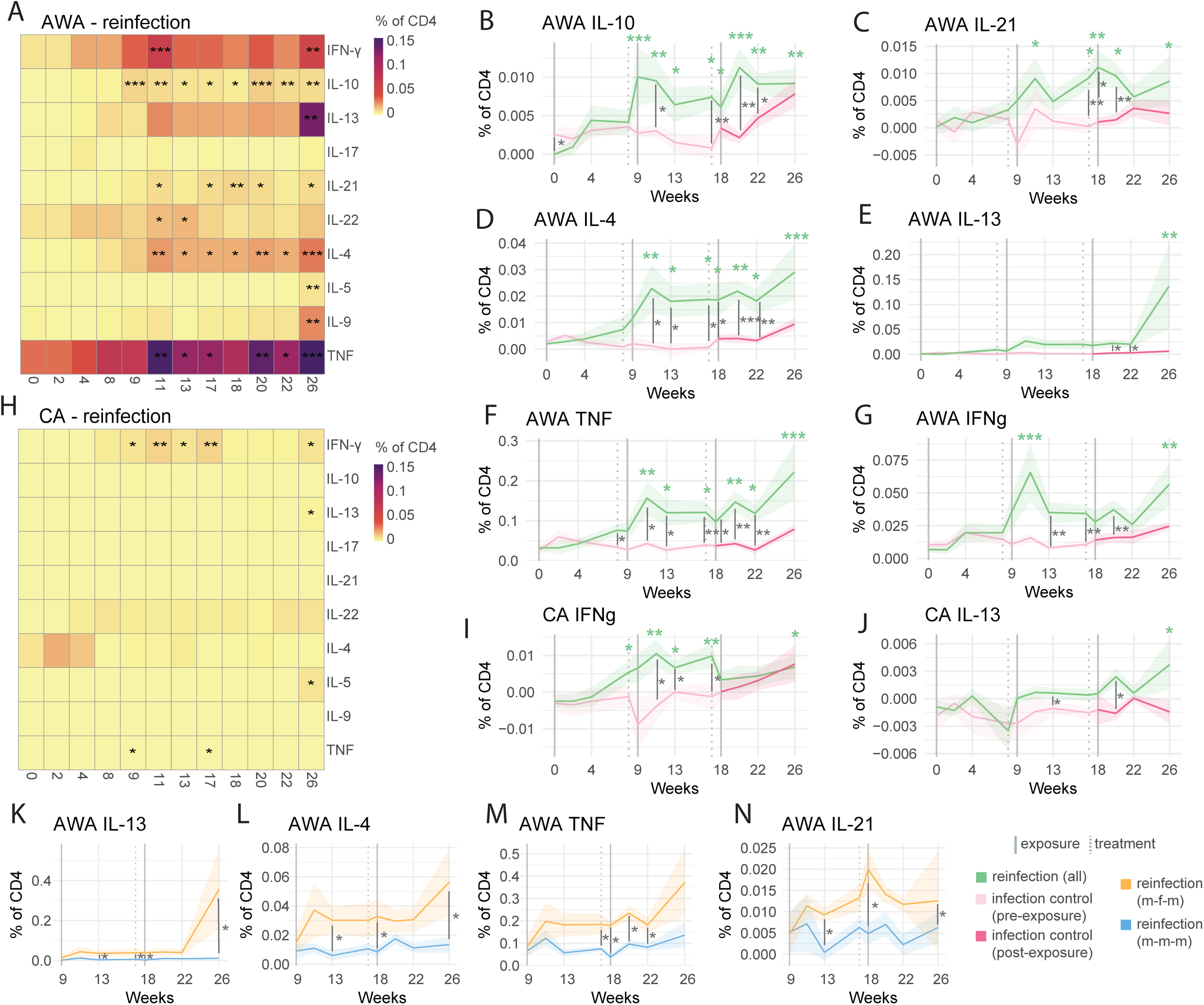
Robust worm (AWA) but not cercariae (CA) - specific CD4 T cell responses during repeat schistosome infection. A) Heatmap displaying frequencies of cytokine positive CD4+ T cells post AWA stimulation of PBMCs from reinfection volunteers, coloured by frequency. B-G) Ribbon plots depicting mean (lines) frequency and standard error (shaded area) of the mean of cytokine production in AWA-stimulated CD4 T cells, coloured by infection group, comparing reinfection (all) and infection control. H) Heatmap displaying frequencies of cytokine positive CD4+ T cells post CA stimulation of PBMCs from reinfection volunteers, coloured by frequency. I-J) Ribbon plots depicting mean (lines) frequency and standard error (shaded area) of the mean of cytokine production in CA-stimulated CD4 T cells, coloured by infection group, comparing reinfection (all) and infection control. In panels A-J a linear mixed model was performed to (separately) assess changes in the reinfection and infection control group compared to the week 0 baseline, with volunteer as a random effect. FDR values are displayed in black (panels A-B) or green (panels C-J) for reinfection (all) or red (infection control) * FDR <0.05, ** FDR<0.01, ***FDR<0.001. To compare between reinfection (all, n=11-12) and infection control groups (n=11) welch’s T-tests were performed at each timepoint with significant p values (uncorrected) displayed with a grey line * p <0.05, ** p <0.01. ** p <0.001. K-M) Ribbon plots depicting mean (lines) frequency and standard error (shaded area) of the mean of specified stimuli and populations, coloured by infection group, comparing reinfection (m-f-m) and reinfection (m-m-m). To compare between reinfection (m-m-m, n=7) and reinfection (m-f-m, n=4-5) groups Mann-Whitney tests were performed at each timepoint with significant p values (uncorrected) displayed with a grey line * p <0.05. All frequencies shown are post medium subtraction.

Within the reinfection group, the m-f-m group had significantly higher AWA-specific IL-13, IL-4, TNF, IL-2, and IFN-y than the m-m-m group from week 13 but most strikingly at week 26 (Fig. 2K-N, Supp. Fig. 2O-X). Cytokine changes were not entirely driven by the m-f-m group, with AWA-specific cytokines increased in the m-m-m group, both compared to baseline week 0 (IL-4, IL-13, IL-10, IFN-γ, TNF) and infection control (IL-4, IL-10, IFN-γ, TNF) (Supp. Fig. 3A-E).

Taken together, repeated CHI elicits a robust and mixed - Th1/Th2/regulatory CD4^+^ T cell-specific cytokine response to AWA, and limited response to cercarial antigen (CA). Schistosome-specific CD4 T cell responses tend to increase after the first infection/treatment cycle (week 11 onwards) and are notably higher in the m-f-m group at week 26.

### Enhanced inflammation due to m-f-m exposure is short-lived, tending to reduce by week 30

Having observed robust alterations in the CD4^+^T cell compartment in the m-f-m repeated infection at week 26, we asked if this response would continue to increase at week 30 (the latest pre-treatment time point) (Fig. 3). We observed that CD38^AND/OR^HLADR^+^ EM T cells increased at week 26 (Fig. 1G), then decreased by week 30 in the m-f-m group, while remaining significantly higher than the m-m-m group (Fig. 3A). A similar pattern was observed with serum cytokines TNF, CCL23 and CCL4, which tended to decrease by week 30 in the m-f-m group (Fig. 3B). No significant changes in IL-18 or CXCL10 were observed (Supp. Fig. 4A).

**Figure 3:**
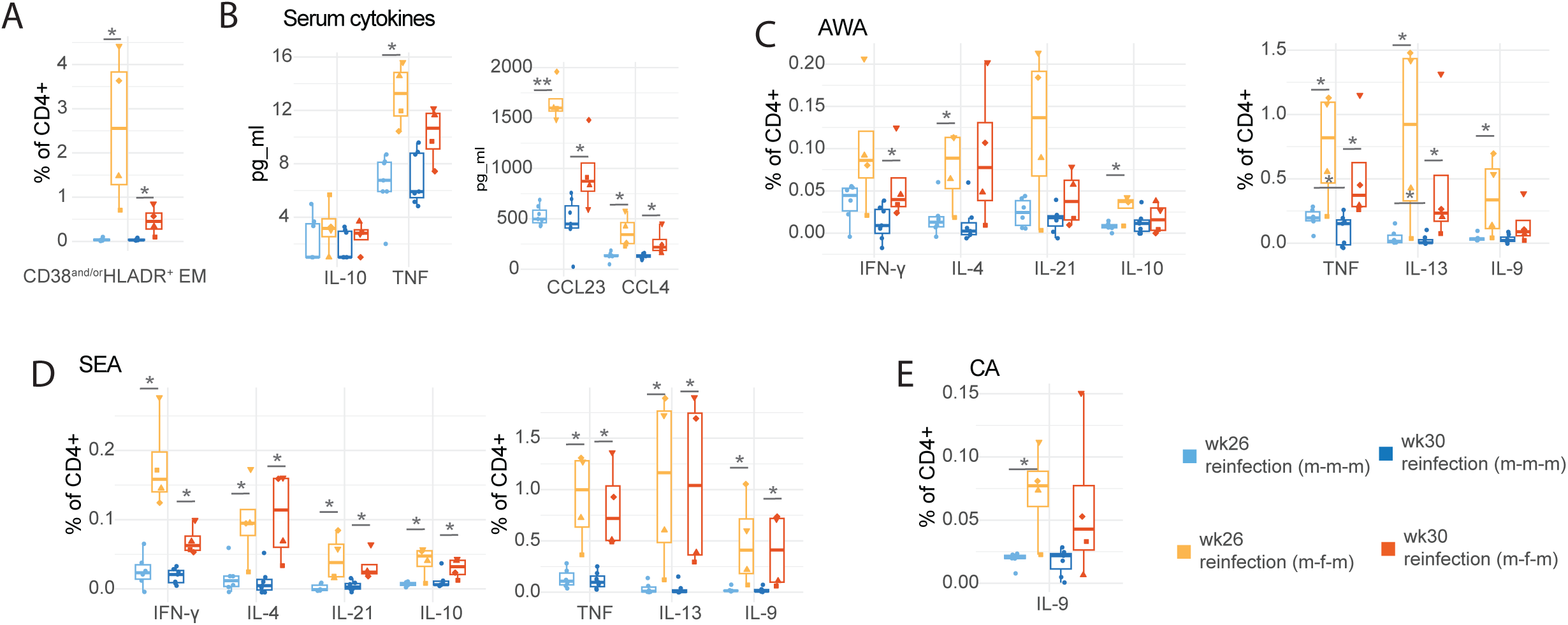
Enhanced inflammation at week 26 in reinfection (m-f-m) group tends to reduce by week 30. A) Boxplot comparing CD38and/orHLADR+ EM CD4 T cell frequency between reinfection (m-m-m) and reinfection (m-f-m) at week 26 and 30. B) Boxplot comparing serum cytokines between reinfection (m-m-m, n=4) and reinfection (m-f-m, n=6) at week 26 and 30. C-E) Boxplots comparing AWA-specific (A), SEA-specific (D) and CA-specific (E) CD4 T cell cytokine frequency between reinfection (m-m-m) and reinfection (m-f-m) at week 26 and 30. All frequencies shown are post medium subtraction. Differing shapes in reinfection (m-f-m) volunteers denote different individuals and are consistent between panels. Boxplots display central line (median), and hinges (25th and 75th percentile) with whiskers extending from the hinge to the largest/smallest value or 1.5x the interquartile range. P values (uncorrected) are displayed, derived from wilcoxon signed-rank tests when comparing between timepoints, or Mann-Whitney test to compare reinfection (m-m-m) and reinfection (m-f-m) groups. * p<0.05, ** p<0.01, ***p<0.001.

Stimulations with adult worm (AWA), egg (SEA), and cercarial (CA) antigens were performed, to understand how schistosome stage-specific responses changed between weeks 26 and 30. We observed robust SEA, AWA (but not CA) responses at week 26 in the m-f-m reinfection group, higher than m-m-m, and encompassing a wide range of Th1/Th2/regulatory cytokines (Fig. 3C-E, Supp. Fig. 4B-D). Cytokine responses to AWA tended to decrease by week 30 (except IL-4) (Fig. 3C). In comparison, and with the notable exception of IFN-y, responses to SEA tended to remain stable to week 30 (Fig. 3D).

### Muted responses in endemic schistosome infection are more comparable with male-male-male (m-m-m) than male-female-male (m-f-m) repeated CHI participants

Next, we explored how T cell and cytokine response in repeated CHI (m-m-m and m-f-m repeated infections) compares to that in natural endemic (mixed-sex) infections. Repeated CHI samples were selected at the last time point before the final PZQ treatment (week 30) when their exposure history most closely echoed endemic populations. Endemic-infected individuals were chosen from a population with regular lake contact (entailing repeated exposure), varying infection burdens, and exposure histories (Supp. Fig. 5). These samples were grouped into “infected” and “uninfected” groups, based on stool Kato-Katz, supported by serum-Circulating Anodic Antigen (CAA) and urine-Circulating Cathodic Antigien (CCA) assays (Supp. Fig. 5). To note, endemic samples had reduced viability - with median viability of 70% - 80% for endemic participants - compared to 96% in the CHI reinfection (all) group (Supp. Fig. 6A). However, we found no evidence that viability was correlated to T cell phenotype or cytokine expression within endemic samples (Supp. Fig. 6).

Using PCA to define an overview of how unstimulated T cell phenotypes differed between m-m-m, m-f-m and natural-endemic-infection groups, we observed that the groups tended to separate along the principal components (Fig. 4A). Activated CD38^AND/OR^HLADR^+^ EM T cells were highest in the reinfection (m-f-m) group (Fig. 4B). Endemic-infected individuals tended to have higher frequencies of co-inhibitory ligand CTLA4 as well as PD1 expressing CD4 T cells though unexpectedly lower Foxp3^+^CD25^+^ Tregs when compared to repeated CHI groups (Fig. 4B). Meanwhile, serum cytokines in endemic infected individuals tended to most closely resemble m-m-m reinfection, although with increased regulatory cytokine IL-10 levels and reduced eosinophil-associated chemokine CCL23 levels (Fig. 4C). The m-f-m infection group had higher serum levels of TNF, CCL23 and CCL4, an effect mainly driven by two of the participants with exaggerated responses in all outputs (Fig. 4C). Notably, endemic serum samples had an overall more anti-inflammatory state, with increased IL-10 to TNF ratio (Fig. 4D).

**Figure 4:**
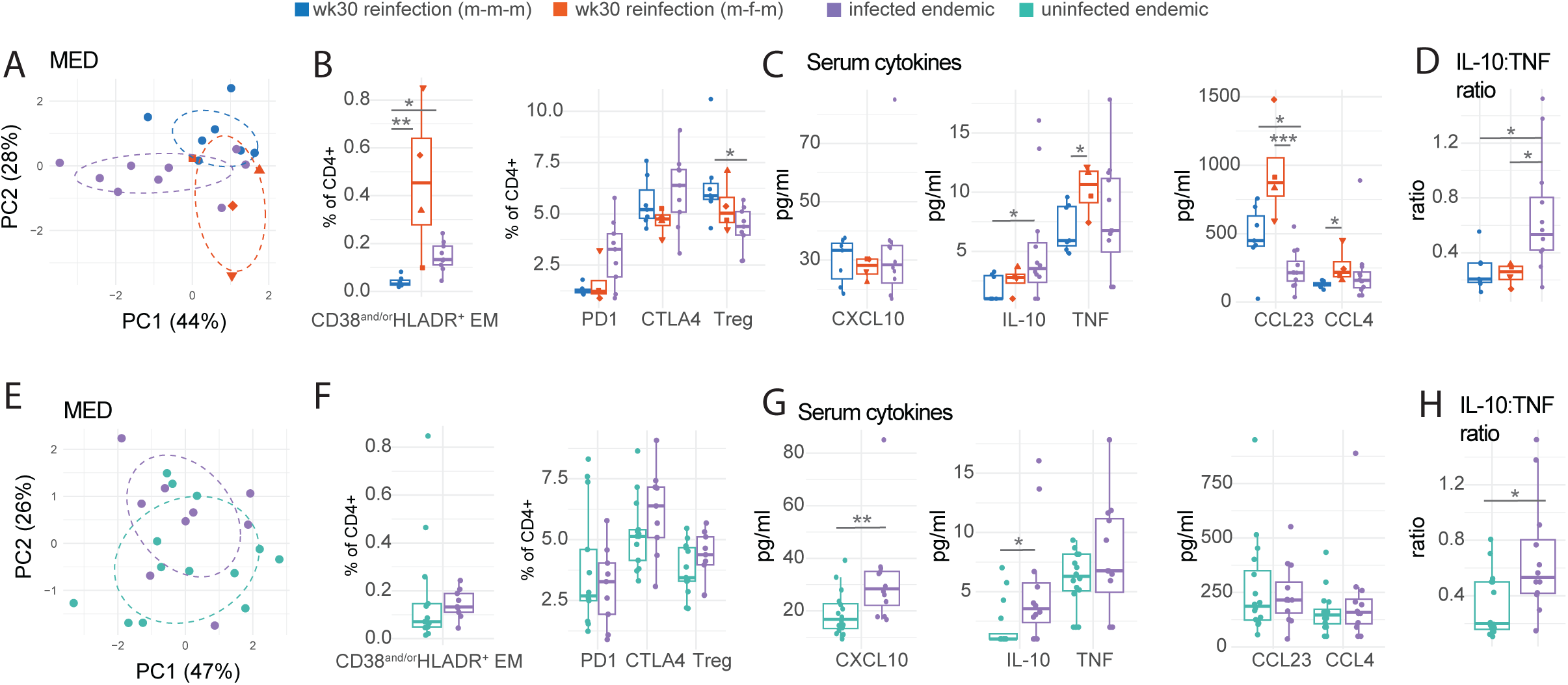
Comparing T cell phenotype and serum cytokines in controlled human infection and endemic infection. A-C) Comparisons of reinfection (m-m-m, n = 7), reinfection (m-f-m, n = 4) and endemic schistosome infected individuals (n=9). Samples are coloured by group. A) PCA applied to individual samples from on the basis of frequencies of T cell populations. B) Boxplots comparing T cell phenotype. C) Boxplots comparing serum cytokine levels. D) Boxplot comparing serum IL10:TNF ratio. E-G) Comparisons of uninfected endemic (n=13) and schistosome infected endemic (n=9) individuals. Samples are coloured by group. E) PCA applied to individual samples from on the basis of frequencies of T cell populations. F) Boxplots comparing T cell phenotype. G) Boxplots comparing serum cytokine levels. H) Boxplot comparing serum IL10:TNF ratio. Boxplots display central line (median), and hinges (25th and 75th percentile) with whiskers extending from the hinge to the largest/smallest value or 1.5x the interquartile range. P values are displayed, these are derived from Dunn’s test for pairwise multiple comparisons in B-D and from a Mann-Whitney test in E-H. * p<0.05, ** p<0.01, *** p<0.001.

We also compared the observed responses in endemic-infected and endemic-uninfected individuals. No significant differences in T cell phenotypes were observed in infected and non-infected participants, potentially attributable to the heterogeneity in these populations (Fig. 4E-G). However, serum cytokines including CXCL10 and IL-10 were significantly elevated in the infected endemic individuals compared to their uninfected counterparts (Fig. 4G). This increase in CXCL10 has been previously reported in this sample set (*14*), however, our study extends these findings by demonstrating that the CXCL10 level observed is comparable to that seen after repeated CHI (Fig. 4C) (*14, 18, 23*). No differences in TNF, CCL23, or CCL4 serum levels were observed (Fig. 4G). In endemic samples, schistosome infection leads to a more anti-inflammatory state, with an increased IL-10 to TNF ratio (Fig. 4H).

Next, we compared schistosome stage-specific responses in the repeated CHI and endemic infection groups. PCA of individual samples post-stimulation revealed a distinct clustering of the SEA-stimulated reinfection m-f-m group, seen to a lesser extent in AWA-stimulation and not observed following CA-stimulation (Fig.5A, D, G). This observation could be indicative of an acute response to SEA due to potential priming by eggs in the reinfection m-f-m group. Considering specific CD4^+^ T cell cytokines, SEA and AWA antigens elicited the highest response, with CA-specific responses the least. AWA and SEA-specific Th1/Th2/regulatory cytokine levels were highest in the reinfection m-f-m group, significant for SEA-specific TNF, IL-13, IL-9, IFN-y, IL-4, IL-21, IL-10 and AWA-specific TNF, IL-13 and IFN-y (Fig. 5B, E). Interestingly, single-sex (m-m-m reinfection) and natural endemic infection elicited comparable antigen-specific cytokine responses, with a notable exception of IL-17 which was higher in endemic infection (Fig.5 B, E, and H). When comparing endemic-infected and uninfected participants, specific responses were observed to AWA and CA (Fig. 5C, I), with close to an absent response to SEA (Fig. 5F). Infected endemic groups exhibited significantly higher levels of AWA-specific IL-4 and IL-10 and CA-specific TNF, IL-9, and IL-17 in comparison to uninfected endemic subjects (Fig. 5C and I). No significant changes in IL-5 and IL-22 were observed (Supp. Fig. 4F-H).

**Figure 5:**
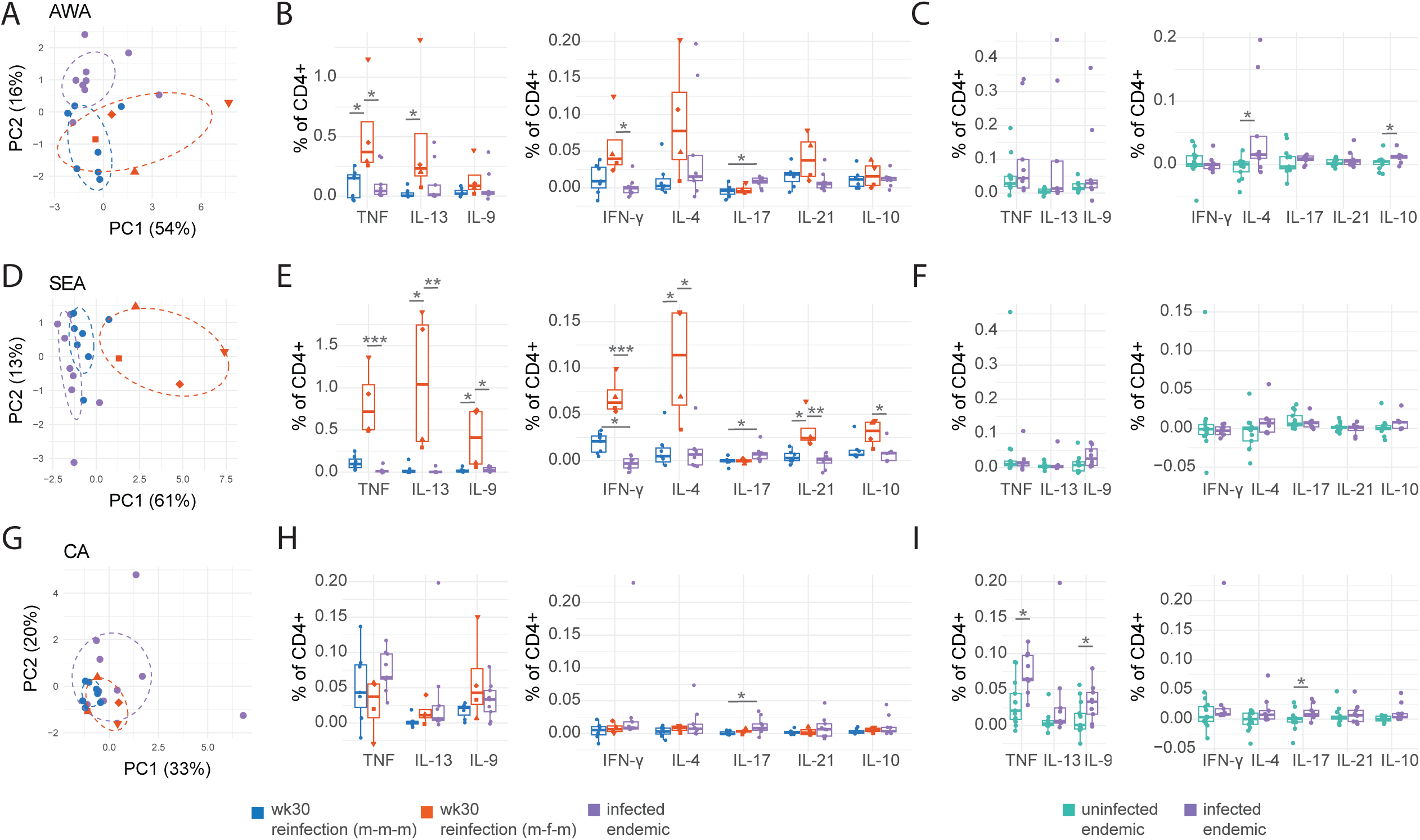
Responses to different schistosome stages (AWA, SEA, CA) in controlled human infection and endemic infection. A, B, D, E, G, H) Comparisons of reinfection (m-m-m, n = 6), reinfection (m-f-m, n = 4) and endemic schistosome infected individuals (n=11). Samples are coloured by group. C, F, I) Comparisons of uninfected (n=13) and schistosome infected (n=9) endemic individuals. Samples are coloured by group. A) PCA applied to individual samples on the basis of AWA-specific cytokine frequencies, coloured by group. B & C) Boxplots comparing AWA-specific CD4 T cell cytokine frequencies, coloured by group. D) PCA applied to individual samples on the basis of SEA-specific cytokine frequencies, coloured by group. E & F) Boxplots comparing SEA-specific CD4 T cell cytokine frequencies, coloured by group. G) PCA applied to individual samples on the basis of CA-specific cytokine frequencies, coloured by group. H & I) Boxplots comparing CA-specific CD4 T cell cytokine frequencies, coloured by group. All boxplots display central line (median), and hinges (25th and 75th percentile) with whiskers extending from the hinge to the largest/smallest value or 1.5x the interquartile range. All frequencies shown are post medium subtraction. P values are displayed, these are derived from Dunn’s test for pairwise multiple comparisons in B, E and H, and from a Mann-Whitney test in C, F and I. * p<0.05, ** p<0.01, *** p<0.001.

Taken together, the immune response in endemic infection is most comparable to that elicited after repeated (m-m-m) CHI, especially responses to worm antigens. No responses to SEA were observed in endemic participants, while CA-specific Th17 response was elevated. Participants in the m-f-m group, with potential egg exposure, showed the highest cytokine response to both AWA and SEA.

## Discussion

This study has delineated how T cell and cytokine responses develop during repeated *S. mansoni* reinfection and treatment cycles; a study design that resembles exposure patterns seen in schistosomiasis-endemic areas. *Schistosoma*-reinfection is characterized by expanded CTLA4^+^ Tregs, CD38^+^CD27^+^DN T cells, and, in the m-f-m group, CD38^AND/OR^HLDR^+^ effector memory expansion. An increase in schistosome worm-specific CD4^+^ T cell cytokine levels consisting of a mixed Th1/Th2/Treg cytokine profile was seen in reinfection (m-m-m) participants. Before final treatment (week 30) AWA-specific responses observed in the m-m-m CHI reinfection group were of a similar magnitude to those in endemic infected participants. Notably, endemic infected individuals showed an anti-inflammatory profile, with a negligible response to SEA and elevated Th17 response to CA. Reinfection m-f-m participants had the highest cytokine responses at weeks 26-30, possibly reflective of an acute egg response (*3, 15, 16*).

Changes in T cell phenotype during m-m-m reinfection were characterized by increased CTLA4^+^ Tregs, potentially contributing to a downregulated host-immune activation, increased parasite survival (*24–26*) and the reduced symptoms as observed during reinfection (*17*). Treg expansion in endemic schistosome infection has previously been described(*27*). In addition, another potentially regulatory T cell subset, CD38^+^CD27^+^DN T cells, expanded at week 8-9, after the first infection, as in the previous CHI (single-sex 1x infection) (*14*). Besides changes in the regulatory T cell phenotype, we also observed a decline in naïve T cells at 11 weeks post-exposure. This declining naïve population post-PZQ treatment could be attributed to immune maturation after antigen release from dying worms (*19, 21*).

In terms of cytokine response, CD4^+^ T cell-specific cytokine responses in m-m-m reinfection were elevated during the first infection cycle, becoming significant after the second exposure. Interestingly, this cytokine response comprised of a mixed-array of Th1/Th2/regulatory cytokines; IFN-γ, TNF, IL-4, IL-13, IL-21 and IL-10, contrary to the long-held view of polarized initial Th1 responses that switch to Th2 on egg production (*28*). We also noted that a significant magnitude of cytokine response was directed to AWA in m-m-m reinfection and a contrastingly negligible CA response. This finding supports the notion that viable cercariae, such as those used in CHIs, rapidly transverse the skin tissue and induce regulatory factors ensuring minimal priming of the host response (*29, 30*).

Compared to the m-m-m group, m-f-m volunteers showed elevated T cell responses, particularly from week 26. Due to reduced PZQ-sensitivity of female worms (*18, 31*), male-female pairing may have occurred in the m-f-m volunteers following the third exposure. Immune differences between the m-m-m and m-f-m appeared from week 26, a time point that coincides with potential egg production (*32*). The elevated Th2 and Th1 responses to egg and worm antigens at this timepoint may therefore reflect an acute response to potential egg presence, a well-characterized phenomenon in the murine model (*33*). Egg presence was not confirmed by egg microscopy or PCR in most of these volunteers, potentially due to insufficient sensitivity to detect low-levels of eggs. Activated CD38^AND/OR^HLA-DR^+^ EM T cells were significantly increased in the m-f-m reinfection group. CD38 and HLA-DR are markers of activation and proliferation expressed by recently activated memory T cells and are often identified in acute inflammatory responses (*34–36*). In this study, the observed T cell activation underscored elevated pro-inflammatory serum chemokines and a cytokine response consisting of CCL23, CCL4, and TNF, further supporting the indication of an acute inflammatory response to egg antigens.

Next, we explored if repeated CHI in non-endemic (Dutch) participants induced immune responses reflective of naturally infected endemic (Ugandan) populations. Naturally infected Ugandan participants showed reduced inflammation, with a tendency for higher co-inhibitory markers (PD1^+^ and CTLA4^+^) and a regulatory serum cytokine environment (elevated IL-10:TNF ratio) compared to repeated CHI participants. This anti-inflammatory phenotype in endemic areas may be attributable to several factors (diet, pathogen exposure, microbiota), and has been associated with a dampened vaccine response (*37, 38*). In line with this, endemic infected participants showed a negligible egg-specific response, a finding in line with previous studies (*39–42*). This dampened egg response may develop as a protective mechanism against egg-induced host tissue damage (*43, 44*). AWA-specific responses in the endemic infected participants were of comparable magnitude to reinfection (m-m-m) CHI volunteers at week 30, though less than that seen in reinfection (m-f-m) volunteers. Finally, an IL-17 response to CA was elevated in endemic infected participants, potentially representing a protective barrier response, as this cytokine has been linked with protection against infection in murine models (*45*).

Utilizing advanced cytometry techniques and a CHI model with the inherent advantage of a defined schistosome cercariae infection dose, sex, and timing has enabled us to conduct a longitudinal assessment of immune changes to reinfection over time. No volunteers developed resistance to infection (*17*), suggesting that the mixed Th1/Th2 response we observed is not sufficient for protection and that vaccines should aim to induce a different T cell phenotype, magnitude or specificity. Our findings are in line with the “happy valley hypothesis”, which argues that whilst extreme Th1 or Th2 responses can be protective, the balanced Th1/Th2 response we observe here may allow for worm survival (*46*). It is possible that developing protective immunity may require years of exposure/reinfection and treatment cycles, with the shorter duration of CHI models presenting a potential limitation. Furthermore, we show how immune responses can develop during repeated infection/treatment cycles using a CHI model, with worm-specific responses after repeated m-m-m CHI immune resembling those of naturally infected endemic participants. Transferring the CHI model to an endemic area is a crucial next step for a holistic immune assessment that accounts for host-specific factors that may influence immune response dynamics during schistosome (re)infection in endemic populations.

## Methods

### Study design

This study utilized samples from a repeated controlled human challenge infection (CHI) model (NCT05085470), a placebo-controlled randomized trial conducted at the Leiden University Medical Centre (LUMC), The Netherlands (*17*) and the HALLMARK study conducted at Uganda Virus Research Institute (UVRI), Entebbe, Uganda (*47*). In the repeated CHI trial, participants in the reinfection group (n=12) were exposed to 20 cercariae at three time points; weeks 0, 9, and 18), with one group receiving male-only cercariae at all time points (m-m-m group, n=7) and the other (n=5) received a sequential exposure to cercariae of different sexes (m-f-m) at the three exposure timepoints, respectively. One participant in the reinfection m-f-m group was lost to follow-up shortly after the third exposure. The infection control group (n=12) received two mock challenges at weeks 0 and 9, and one 20 male-only cercariae dose exposure at week 18. Praziquantel (PZQ) treatment was administered 8 weeks following the first and second exposures (weeks 8 and 17) and at 12 weeks after the third challenge (week 30) with 60mg/kg PZQ or placebo (*17*). *Schistosoma*-specific PCR was performed on stool samples, as previously described (*48*) to detect the presence of schistosome eggs. Endemic human samples were obtained from 18-25 year old young adults living in the schistosomiasis-endemic area of Kigungu landing site in Entebbe municipality, Uganda (*47*). This was a cross-sectional study consisting of volunteers with no active infection at the point of enrolment (endemic-uninfected group) and those with active infection (endemic-infected group).

### Determination of current and prior infection

In the repeated CHI trial, volunteer infection status was determined using 500 µl of serum and the upconverting nano-particle lateral-flow CAA assay at a cut-off point ≥1 pg/ml (*49*). Meanwhile, infection status in the endemic study was determined by Kato Katz, supported by serum CAA and urine CCA (Supp. Figure 5). Serum CAA in the endemic study was detected using 20 µl of serum and the upconverting nano-particle lateral flow (UCP-LF CAA) assay at a cut-off point ≥10 pg/ml (the lower sensitivity reflects reduced available sample volume) (*49*). Kato-Katz and urine CCA (Rapid Medical Diagnostics) were measured at the point of sample collection (*49–51*). To ascertain the prior exposure status of Ugandan volunteers, schistosome-specific AWA-specific IgG and SEA-specific IgG were measured using in-house IFA and ELISA assays, respectively (*23, 52*).

### PBMC isolation, storage, and thawing

PBMC isolation from blood samples obtained from endemic infection was performed at UVRI, Uganda, while those from the ReCoHSI study were isolated in LUMC, Netherlands both using the Ficoll gradient density separation method, as previously described (*14*).

In summary, venous whole blood samples collected in heparin tubes were diluted in HBBS (Invitrogen) and then separated over a Ficoll gradient (Apotheek LUMC) by centrifugation at low break for at 400g for 25 minutes at room temperature. PBMCs collected were then washed with HBSS, counted, and cryopreserved in complete RPMI (Invitrogen), with 100 U/ml penicillin G sodium, 100 µg/ml streptomycin (Sigma), 1 mM pyruvate (Sigma), and 2 mM glutamine (Sigma); with 10% DMSO (Merck) and 20% FCS (Bodinco). Cryopreserved cells were stored in a -80°C freezer overnight and transferred to liquid nitrogen for long-term storage. PBMC isolation and cryopreservation at UVRI followed the same protocol except for the following differences: all washes were in complete RPMI, centrifugation by Ficoll gradient was at 1000g for 22 minutes, and cryopreservation used a higher FCS percentage (50%).

Cryopreserved PBMC samples from Uganda were transported to the LUMC and thawed together with frozen PBMCs from the ReCoHSI trial. Cryopreserved PBMC samples were thawed at 37°C in thawing media (complete RPMI 1640, 20% FCS, benzonase 25 units/ml (Merck).

### Stimulation for cytokine production

Up to 1x10^6^ cells per condition were plated into culture media (complete RPMI and 10% FCS) in round-bottom well plates (BD Biosciences) and stimulated with culture media, SEA (10 μg/ml), CA (50 μg/ml) and AWA (50 μg/ml) for 24hrs at 37°C incubation. Frozen (eggs, cercariae) or freeze-dried (adult worms) were used to prepare antigens via homogenization with a glass homogenizer on ice in PBS, followed by sonification. Antigen preparations were frozen overnight at -80°C, thawed, and centrifuged for 25 minutes at 13,000 rpm at 4°C. The antigen preparation supernatants were filter sterilized and their concentration was measured via the bicinchoninic acid assay (Pierce). As a positive control SEB (200 ng/ml) was added to pooled samples. Four hours after the beginning of the incubation, Brefeldin A (5mg/ml, Sigma) was added to wells and then incubated for a further 20 hours. Upon completion of stimulation, cells were centrifuged (at 400xg, for 4 min) in a V-bottom plate (BD Biosciences) and the supernatant transferred to 96-well round-bottomed plates (BD Biosciences) for subsequent cytokine analysis. Two (uninfected) endemic samples and four (infected) endemic samples were excluded due to low viability post-stimulation (<55% live) (Supp. Fig. 6). One CHI participant in the infection control group was excluded from the analysis of schistosome-specific cytokines due to high cytokine production in the unstimulated control (>1%).

### Staining, flow cytometry, and analysis

Before extra- and intra-cellular staining, stimulated cells were washed twice with PBS and stained with live/dead (Blue, Thermofisher, 1:1000) in PBS before a third wash with FACS buffer. Extracellular staining was performed using the following antibodies; CD38 (APC-Fire, Biolegend, 1:1500, 356643), CD8 (Pacific orange, Thermofisher, 1:1000, MHCDO830), CD25 (BUV563, BD, 1:750, 612918), CD27(APC-H7, BD, 1:500, 560222), CD11b (BV510, Biolegend, 1:750, 101263), CD123 (BV510, Biolegend, 1:750, 306022), CD19 (BV605, Biolegend, 1:750, 302244), CD4 (cFlourYG584, CYTEK, 1:750, SKU R7-200), CD3 (BUV395, BD, 1:200,563546), CD27 (APC-H7, BD, 1:500, 560222), CD56 (BV510, Biolegend, 1:500, 318340), PD1 (BV750, BD, 1:375, 747446), γδTCR (BV480, BD, 1:250, 747446), CD127 (R718, BD, 1:100, 566967), CCR7 (BV785, Biolegend, 1:40, 353230), HLA-DR (PE-fire, Biolegend, 1:400, 307683) and human Fc block (Invitrogen, 1:200, 14-91613) to prevent non-specific antibody binding.

Intracellular staining of cells comprised of an overnight incubation with TNF (PE-Cy7, BD, 1:3000, 557647), IL-17A (Pacific blue, Biolegend, 1:3000, 512312), IL-5 (APC, Biolegend, 1:1000, 504306), IL-4 (BUV737, BD, 1: 750, 612835), IFN-γ (BV650, BD, 1:750, 563416), IL-13 (BV711, BD, 1:750, 564288), IL-9 (PE, Biolegend, 1:500, 507605), IL-10 (PerCP ef710, Thermofisher, 1:250, 46-7108-4), CTLA (PE-Cy5, BD, 1: 10000, 555854), IL-21 (AF647, BD, 1:200, 560493), FoxP3 (PE-Dazzle594, Biolegend, 1:150, 320126), IL-22 (Vio515, Miltenyi, 1:100, 130-108-09), GATA3 (BV421, BD, 1:200, 563349), CD45RA (BUV496, BD, 1:750, 741182), CD45RA (BUV496, BD, 1:750, 741182), CD45RO (BUV805, BD, 1:750, 748367) and CRTh2 (BUV661, BD, 1:200, 741663) antibody mix diluted in eBio perm buffer (eBioscience). Stained cells were washed thrice in eBio perm buffer (200µl, eBioscience) and acquired on a spectral flow cytometer (Cytek Aurora). Cytometer output was analyzed in Spectral flow version 3.1 (Cytek) and OMIQ (Dotmatics) software, and gates were positioned using a combination of fluorescence minus many (FMM) controls and medium as negative controls.

### Measurement for serum cytokines

The following cytokines: CCL4, CCL23, CXCL10, IL-5, IL-13, IL-10, IL-18, TNF, and IFN-γ were measured in serum and cell supernatant using a commercial Luminex kit (Cat. Number: LXSAHM, R&D systems), according to manufacturer’s instructions. Cytokines of which over 60% of samples were below the limit of detection were excluded from analysis. For serum cytokines, these were IL-5, IL-13, and IFN-γ. For supernatant cytokines these were IL-5, CCL23, IFN-γ, IL-18.

### Statistical analysis

Statistical analysis and visualization was done in R (v4.3.1 core team 2023) and R studio (version 2023.06.1) (*53*). Response in ReCoHSI infection control- and reinfection groups over time was assessed using a linear mixed model (R package lmerTest v3.1-3) (*54*) with timepoint as a factor and fixed effect, and participant as a random effect. The frequency of T cell subsets and cytokine-positive CD4+ T cells were used as response variables during the analysis to determine T cell and cytokine response outcomes, respectively. P values were FDR-corrected for multiple comparisons. To compare between reinfection (all) and infection control groups Welch’s T-tests were performed at each time point. Normality was not assumed for the reinfection m-f-m group or any endemic group. Therefore, in comparing between reinfection (m-m-m) and reinfection (m-f-m) groups Mann-Whitney tests were performed at each timepoint. To compare immune responses between individual reinfection volunteers at weeks 26 and 30 a Wilcoxon signed-rank test was used (package rstatix v 0.7.2) (*55*). To compare between endemic infected and uninfected a Mann-Whitney test was used, with a Kruskal-Wallis test, followed by the post-hoc Dunn’s test (package rstatix v 0.7.2) (*55*) used to compare to reinfection groups. For all cross-group comparisons, the r package rstatix was used.

## Data Availability

All data produced in the present work are contained in the manuscript. Additional data is available upon reasonable request to the author.

## Acknowledgments

The authors thank the LUMC Klinisch Microbiologisch Laboratorium (KML) for performing antibody diagnostics in this work. Most of all, we thank all the study volunteers for their participation.

## Funding

This research has received funding from the European Union under ERC St grant agreement No 101075876. Part of the work was conducted at the MRC/UVRI and LSHTM Uganda Research Unit which is jointly funded by the UK Medical Research Council (MRC) part of UK Research and Innovation (UKRI) and the UK Foreign, Commonwealth and Development Office (FCDO) under the MRC/FCDO Concordat agreement and is also part of the EDCTP2 programme supported by the European Union. Harriet Mpairwe received a Wellcome Training fellowship 102512, ELH was supported by the European Union’s Horizon 2020 research and innovation program under the Marie Skłodowska-Curie grant (101063914), and ED was supported by a Wellcome grant for ‘Human infection studies for Schistosoma mansoni vaccine testing in Uganda’ (218454z/19/z).

## Author contributions

M. Y., M. R., E. L. H., E. D., J. P. R. K., H. M., A. S. M., M. E. and A. M. E. were responsible for study conceptualization and design. E. D., K. A. S., F. S. and E. L. H. were responsible for the statistical analysis and data interpretation and prepared the first draft. M. Y., M. R., A. M. E., H. M. and A. S. M. acquired funding. J. P. R. K. and M. R. were the clinical investigators. J. J. J. performed the trial management. M. C-P., J. S., and J. J. J. were responsible for the production and release of cercariae. F. S., M. K., Y.K., S. T. H, S. S., H. d. B-R., E. I., E. D. and E. L. H. generated the data and optimized the experimental protocols. H. d. B-R., E. I., I. N., M. K. and Y. K. coordinated PBMC sample collection and processing. S. T. H., G. J. v. D., P. L. A. M. C and L. v. L. supported diagnostic (CAA, IFA and ELIZA) tests. All authors contributed to manuscript review.

## Competing interests

The authors declare that they have no conflict of interest. Funding sources did not play any role in the data collection, analyzing, interpretation and or data reporting. Opinions expressed in this paper reflect only the authors’ views and funding sources are not responsible for the use that maybe made of and from the contents of this document.

## Notes

### Competing Interest Statement

The authors have declared no competing interest.

### Clinical Trial

NCT05085470

### Funding Statement

This research received funding from the European Union. Part of the work was conducted at the MRC/UVRI and LSHTM Uganda Research Unit which is jointly funded by the UK Medical Research Council part of UK Research and Innovation and the UK Foreign, Commonwealth and Development Office.

### Author Declarations

This study received ethical approval from De Medical Ethics Committee Leiden The Hague Delft, Netherlands. Part of the work on endemic setting received ethical approval from the Ugandan Virus Research Institue Research Ethics committee and regulatory approval was obtained from the Ugandan National Council for Science and Technology

## References

1. X.-Y. Wang et al., Prevalence and correlations of schistosomiasis mansoni and schistosomiasis haematobium among humans and intermediate snail hosts: a systematic review and meta-analysis. Infectious Diseases of Poverty 13, 63 (2024).

2. A. H. Costain, A. S. MacDonald, H. H. Smits, Schistosome Egg Migration: Mechanisms, Pathogenesis and Host Immune Responses. Frontiers in Immunology 9, (2018).

3. E. J. Pearce, P. Caspar, J. M. Grzych, F. A. Lewis, A. Sher, Downregulation of Th1 cytokine production accompanies induction of Th2 responses by a parasitic helminth, Schistosoma mansoni. J Exp Med 173, 159–166 (1991).

4. A. J. Molehin, Current Understanding of Immunity Against Schistosomiasis: Impact on Vaccine and Drug Development. Res Rep Trop Med 11, 119–128 (2020).

5. M. M. Chaponda, H. Y. P. Lam, Schistosoma antigens: A future clinical magic bullet for autoimmune diseases? Parasite 31, 68 (2024).

6. R. S. Barsoum, G. Esmat, T. El-Baz, Human Schistosomiasis: Clinical Perspective: Review. Journal of Advanced Research 4, 433–444 (2013).

7. T. Hailegebriel, E. Nibret, A. Munshea, Efficacy of Praziquantel for the Treatment of Human Schistosomiasis in Ethiopia: A Systematic Review and Meta-Analysis. J Trop Med 2021, 2625255 (2021).

8. A. Nalugwa, F. Nuwaha, E. M. Tukahebwa, A. Olsen, Single Versus Double Dose Praziquantel Comparison on Efficacy and Schistosoma mansoni Re-Infection in Preschool-Age Children in Uganda: A Randomized Controlled Trial. PLoS Negl Trop Dis 9, e0003796 (2015).

9. C. L. Black et al., Influence of exposure history on the immunology and development of resistance to human Schistosomiasis mansoni. PLoS Negl Trop Dis 4, e637 (2010).

10. G. C. Milne, et al., Revisiting immunity vs. exposure in schistosomiasis: A mathematical modeling study of delayed concomitant immunity. PNAS Nexus 3, pgae471 (2024).

11. L. M. Ganley-Leal et al., Correlation between eosinophils and protection against reinfection with Schistosoma mansoni and the effect of human immunodeficiency virus type 1 coinfection in humans. Infect Immun 74, 2169–2176 (2006).

12. R. R. Oliveira et al., Factors associated with resistance to Schistosoma mansoni infection in an endemic area of Bahia, Brazil. Am J Trop Med Hyg 86, 296–305 (2012).

13. E. C. Mbanefo et al., Host Determinants of Reinfection with Schistosomes in Humans: A Systematic Review and Meta-analysis. PLOS Neglected Tropical Diseases 8, e3164 (2014).

14. E. L. Houlder, et al., Early symptom-associated inflammatory responses shift to type 2 responses in controlled human schistosome infection. Sci Immunol 9, eadl1965 (2024).

15. J. M. Grzych et al., Egg deposition is the major stimulus for the production of Th2 cytokines in murine schistosomiasis mansoni. The Journal of Immunology 146, 1322–1327 (1991).

16. M. Okano, A. R. Satoskar, K. Nishizaki, M. Abe, D. A. Harn, Jr., Induction of Th2 Responses and IgE Is Largely Due to Carbohydrates Functioning as Adjuvants on Schistosoma mansoni Egg Antigens1. The Journal of Immunology 163, 6712–6717 (1999).

17. J. P. R. Koopman et al., Clinical tolerance but no protective efficacy in a placebo-controlled trial of repeated controlled schistosome infection. J Clin Invest, (2024).

18. J. P. R. Koopman et al., Safety and infectivity of female cercariae in Schistosoma-naïve, healthy participants: a controlled human Schistosoma mansoni infection study. eBioMedicine 97, 104832 (2023).

19. E. Linder, C. Thors, Schistosoma mansoni: praziquantel-induced tegumental lesion exposes actin of surface spines and allows binding of actin depolymerizing factor, gelsolin. Parasitology 105 (**Pt 1**), 71–79 (1992).

20. N. Reimers et al., Drug-induced exposure of Schistosoma mansoni antigens SmCD59a and SmKK7. PLoS Negl Trop Dis 9, e0003593 (2015).

21. S.-H. Xiao, J. Sun, M.-G. Chen, Pharmacological and immunological effects of praziquantel against Schistosoma japonicum: a scoping review of experimental studies. Infectious Diseases of Poverty 7, 9 (2018).

22. C. M. Fitzsimmons et al., Chemotherapy for schistosomiasis in Ugandan fishermen: treatment can cause a rapid increase in interleukin-5 levels in plasma but decreased levels of eosinophilia and worm-specific immunoglobulin E. Infect Immun 72, 4023–4030 (2004).

23. M. C. C. Langenberg et al., A controlled human Schistosoma mansoni infection model to advance novel drugs, vaccines and diagnostics. Nature Medicine 26, 326–332 (2020).

24. C. A. M. Finney, M. D. Taylor, M. S. Wilson, R. M. Maizels, Expansion and activation of CD4+CD25+ regulatory T cells in Heligmosomoides polygyrus infection. European Journal of Immunology 37, 1874–1886 (2007).

25. N. Sobhani et al., CTLA-4 in Regulatory T Cells for Cancer Immunotherapy. Cancers (Basel) 13, (2021).

26. M. P. J. White, C. M. McManus, R. M. Maizels, Regulatory T-cells in helminth infection: induction, function and therapeutic potential. Immunology 160, 248–260 (2020).

27. Y. Schmiedel et al., CD4(+) CD25(hi)FOXP3(+) Regulatory T Cells and Cytokine Responses in Human Schistosomiasis before and after Treatment with Praziquantel. Plos Neglected Tropical Diseases 9, (2015).

28. E. J. Pearce et al., Th2 response polarization during infection with the helminth parasite Schistosoma mansoni. Immunol Rev 201, 117–126 (2004).

29. K. G. Hogg, S. Kumkate, S. Anderson, A. P. Mountford, Interleukin-12 p40 secretion by cutaneous CD11c+ and F4/80+ cells is a major feature of the innate immune response in mice that develop Th1-mediated protective immunity to Schistosoma mansoni. Infect Immun 71, 3563–3571 (2003).

30. A. P. Mountford, F. Trottein, Schistosomes in the skin: a balance between immune priming and regulation. Trends in Parasitology 20, 221–226 (2004).

31. L. Pica-Mattoccia, D. Cioli, Sex- and stage-related sensitivity of Schistosoma mansoni to in vivo and in vitro praziquantel treatment. Int J Parasitol 34, 527–533 (2004).

32. R. F. Sturrock, A. E. Butterworth, V. Houba, Schistosoma mansoni in the baboon (Papio anubis): parasitological responses of Kenyan baboons to different exposures of a local parasite strain. Parasitology 73, 239–252 (1976).

33. J. M. Grzych et al., Egg deposition is the major stimulus for the production of Th2 cytokines in murine schistosomiasis mansoni. J Immunol 146, 1322–1327 (1991).

34. C. Sandoval-Montes, L. Santos-Argumedo, CD38 is expressed selectively during the activation of a subset of mature T cells with reduced proliferation but improved potential to produce cytokines. J Leukoc Biol 77, 513–521 (2005).

35. W. Li, L. Liang, Q. Liao, Y. Li, Y. Zhou, CD38: An important regulator of T cell function. Biomedicine & Pharmacotherapy 153, 113395 (2022).

36. A. Ahmed et al., Circulating HLA-DR+CD4+ effector memory T cells resistant to CCR5 and PD-L1 mediated suppression compromise regulatory T cell function in tuberculosis. PLoS Pathog 14, e1007289 (2018).

37. E. Muyanja et al., Immune activation alters cellular and humoral responses to yellow fever 17D vaccine. J Clin Invest 124, 3147–3158 (2014).

38. M. van Dorst et al., Immunological factors linked to geographical variation in vaccine responses. Nat Rev Immunol 24, 250–263 (2024).

39. A. R. de Jesus et al., Clinical and immunologic evaluation of 31 patients with acute schistosomiasis mansoni. J Infect Dis 185, 98–105 (2002).

40. M. E. Williams et al., Leukocytes of patients with Schistosoma mansoni respond with a Th2 pattern of cytokine production to mitogen or egg antigens but with a Th0 pattern to worm antigens. J Infect Dis 170, 946–954 (1994).

41. J. L. Grogan, P. G. Kremsner, A. M. Deelder, M. Yazdanbakhsh, Antigen-specific proliferation and interferon-gamma and interleukin-5 production are down-regulated during Schistosoma haematobium infection. J Infect Dis 177, 1433–1437 (1998).

42. S. Joseph et al., Cytokine production in whole blood cultures from a fishing community in an area of high endemicity for Schistosoma mansoni in Uganda: the differential effect of parasite worm and egg antigens. Infect Immun 72, 728–734 (2004).

43. S. K. Lundy, N. W. Lukacs, Chronic schistosome infection leads to modulation of granuloma formation and systemic immune suppression. Front Immunol 4, 39 (2013).

44. D. L. Boros, R. P. Pelley, K. S. Warren, Spontaneous modulation of granulomatous hypersensitivity in schistosomiasis mansoni. J Immunol 114, 1437–1441 (1975).

45. J. M. Inclan-Rico, et al., “MrgprA3 neurons selectively control myeloid-derived cytokines for IL-17 dependent cutaneous immunity”. Res Sq, (2023).

46. R. A. Wilson, P. S. Coulson, Strategies for a schistosome vaccine: can we manipulate the immune response effectively? Microbes and Infection 1, 535–543 (1999).

47. E. L. Houlder et al., Pulmonary inflammation promoted by type-2 dendritic cells is a feature of human and murine schistosomiasis. Nature Communications 14, 1863 (2023).

48. L. Meurs et al., Is PCR the Next Reference Standard for the Diagnosis of Schistosoma in Stool? A Comparison with Microscopy in Senegal and Kenya. PLOS Neglected Tropical Diseases 9, e0003959 (2015).

49. P. Corstjens et al., Circulating Anodic Antigen (CAA): A Highly Sensitive Diagnostic Biomarker to Detect Active Schistosoma Infections-Improvement and Use during SCORE. Am J Trop Med Hyg 103, 50–57 (2020).

50. M. Adriko et al., Evaluation of circulating cathodic antigen (CCA) urine-cassette assay as a survey tool for Schistosoma mansoni in different transmission settings within Bugiri District, Uganda. Acta Trop 136, 50–57 (2014).

51. P. L. Corstjens et al., Tools for diagnosis, monitoring and screening of Schistosoma infections utilizing lateral-flow based assays and upconverting phosphor labels. Parasitology 141, 1841–1855 (2014).

52. A. M. Deelder, R. J. M. van Zeyl, Y. E. Fillié, J. P. Rotmans, W. Duchenne, Recognition of gut-associated antigens by immunoglobulin M in the indirect fluorescent antibody test for schistosomiasis mansoni. Transactions of the Royal Society of Tropical Medicine and Hygiene 83, 364–367 (1989).

53. RCoreTeam, in R Foundation for Statistical Computing, Vienna, Austria. (2023).

54. A. Kuznetsova, P. B. Brockhoff, R. H. B. Christensen, lmerTest Package: Tests in Linear Mixed Effects Models. Journal of Statistical Software 82, 1–26 (2017).

55. K. Alboukadel, in R package version 0.7.2. (2023).

