## Supplemental figures for "T cell responses in repeated controlled human schistosome infection compared to natural exposure"

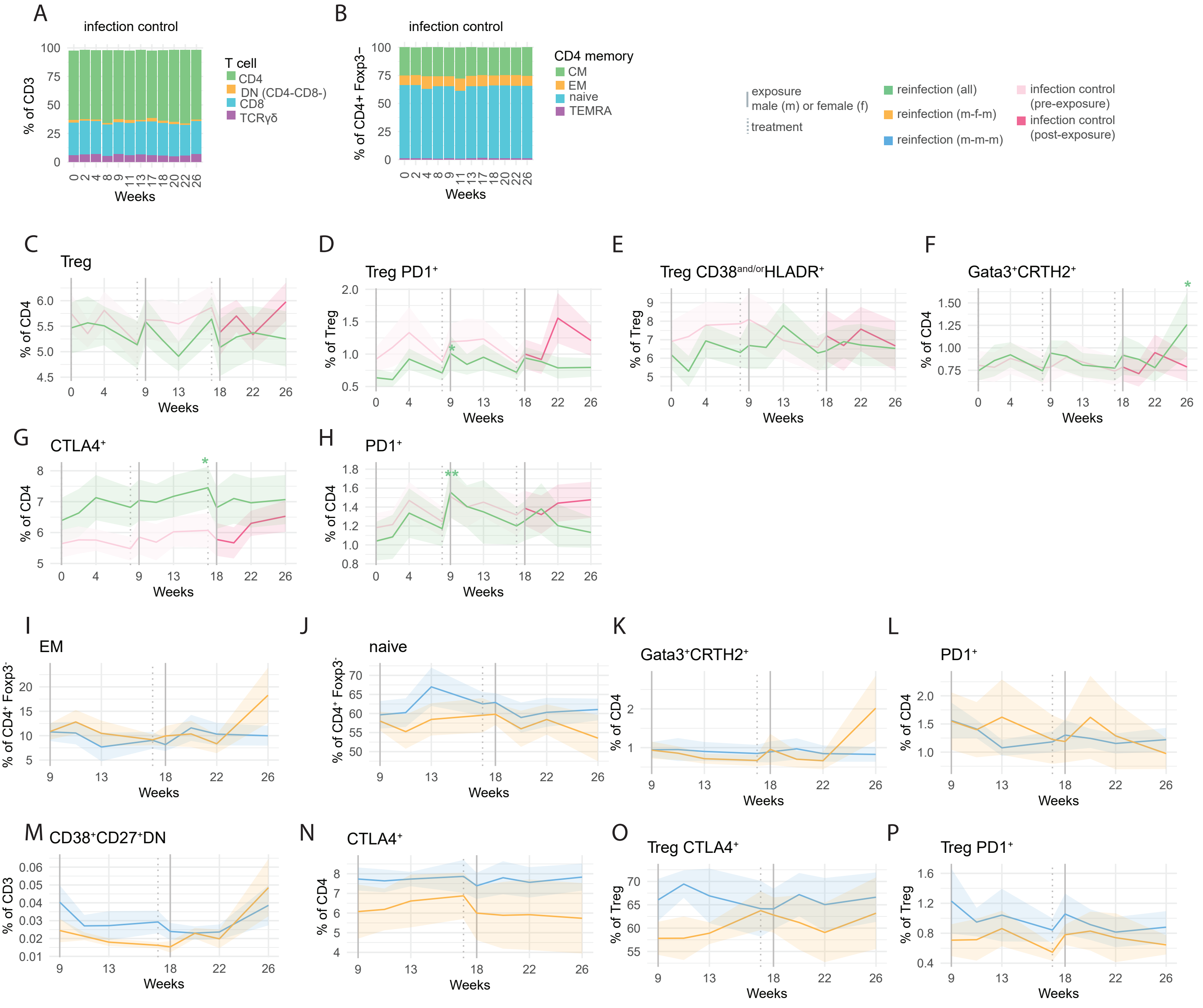


**Supp. Figure 1: Changes in unstimulated T cell phenotype during repeat controlled human schistosome infection.**
A) Stacked bar chart of T cell subset frequencies and B) CD4 T cell memory subset frequencies in infection control group. Each bar section represents the mean frequency. C-H) Ribbon plots depicting mean (lines) frequency and standard error (shaded area) of the mean of specified populations, coloured by infection group. In panels C-H a linear mixed model was performed to (separately) assess changes in the reinfection and infection control group compared to the week 0 baseline, with volunteer as a random effect. FDR values are displayed in black in green (reinfection) and pink (infection control) in figures C-H. * FDR <0.05, ** FDR<0.01. To compare between reinfection (all) and infection control groups welch's T-tests were performed at each timepoint, no significant values were found. I-P) Ribbon plots depicting mean (lines) frequency and standard error (shaded area) of the mean of specified cell populations in the reinfection (m-m-m) and reinfection (m-f-m) groups. To compare between reinfection (m-m-m) and reinfection (m-f-m) groups Mann-Whitney tests were performed at each timepoint, with no significant p values found.

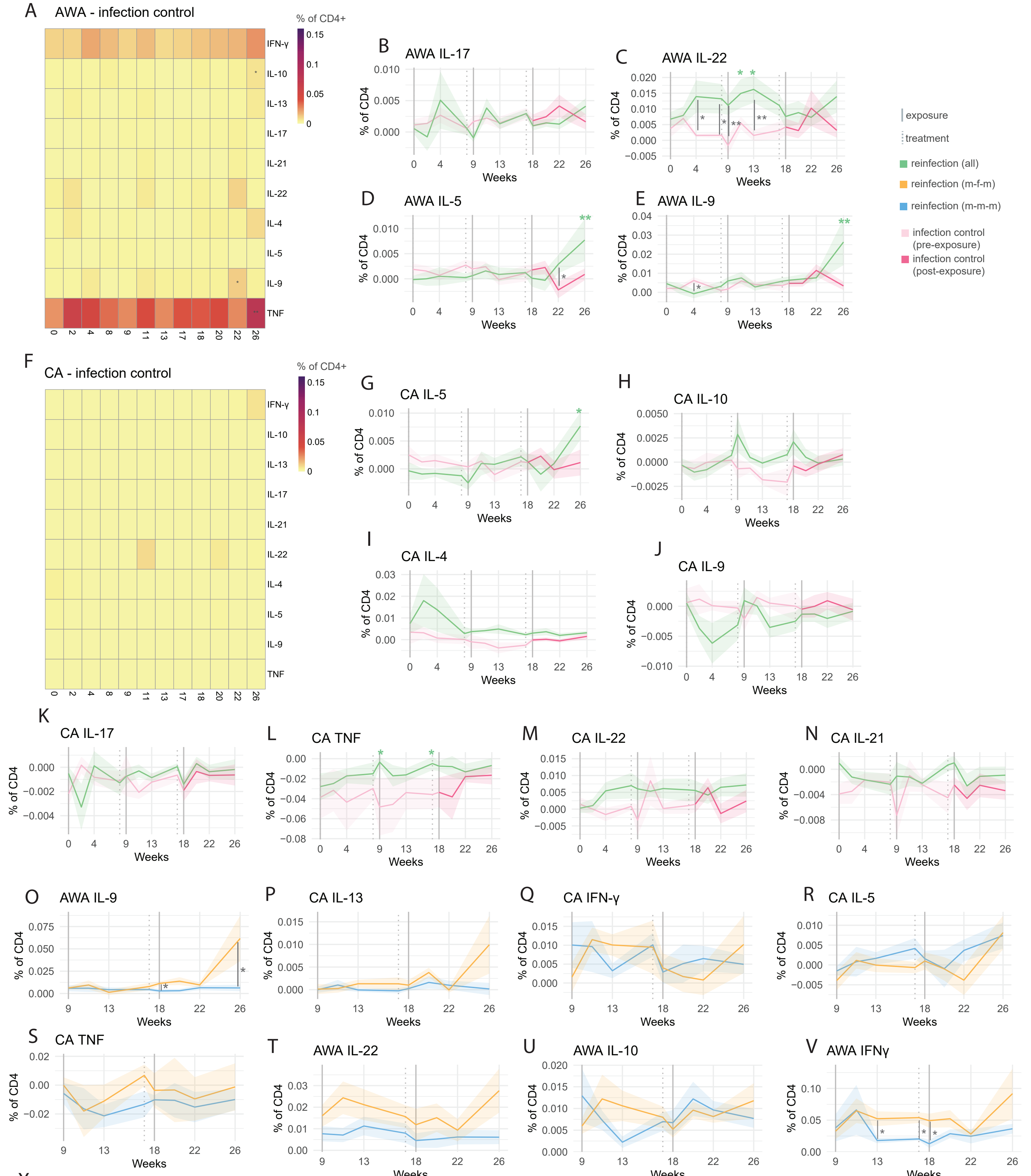

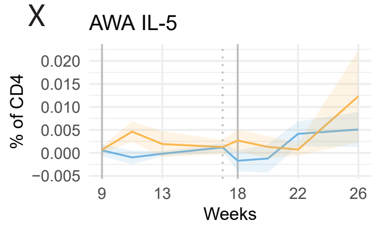


**Supp. Figure 2: Adult worm (AWA) and cercariae (CA) - specific CD4 T cell responses during repeat schistosome infection.**
A,F) Heatmaps displaying frequencies of cytokine positive CD4⁺ T cells post AWA (A) or CA (F) stimulation of PBMCs from infection control volunteers, coloured by frequency. B-E,G-N) Ribbon plots depicting mean (lines) frequency and standard error (shaded area) of the mean of specified stimuli and populations, coloured by infection group, comparing reinfection (all) and infection control. In panels B-E,G-N a linear mixed model was performed to (separately) assess changes in the reinfection and infection control group compared to the week 0 baseline, with volunteer as a random effect. FDR values are displayed in black (panels A,F) or green (panels B-E,G-N) for reinfection (all) or red (infection control) * FDR <0.05, ** FDR<0.01. To compare between reinfection (all) and infection control groups welch's T-tests were performed at each timepoint with significant p values (uncorrected) displayed with a grey line * p <0.05, ** p <0.01. O-X) Ribbon plots depicting mean (lines) frequency and standard error (shaded area) of the mean of specified stimuli and populations, coloured by infection group, comparing reinfection (m-f-m) and reinfection (m-m-m). To compare between reinfection (m-m-m) and reinfection (m-f-m) groups Mann-Whitney tests were performed at each timepoint with significant p values (uncorrected) displayed with a grey line * p <0.05. All frequencies shown are post medium subtraction.

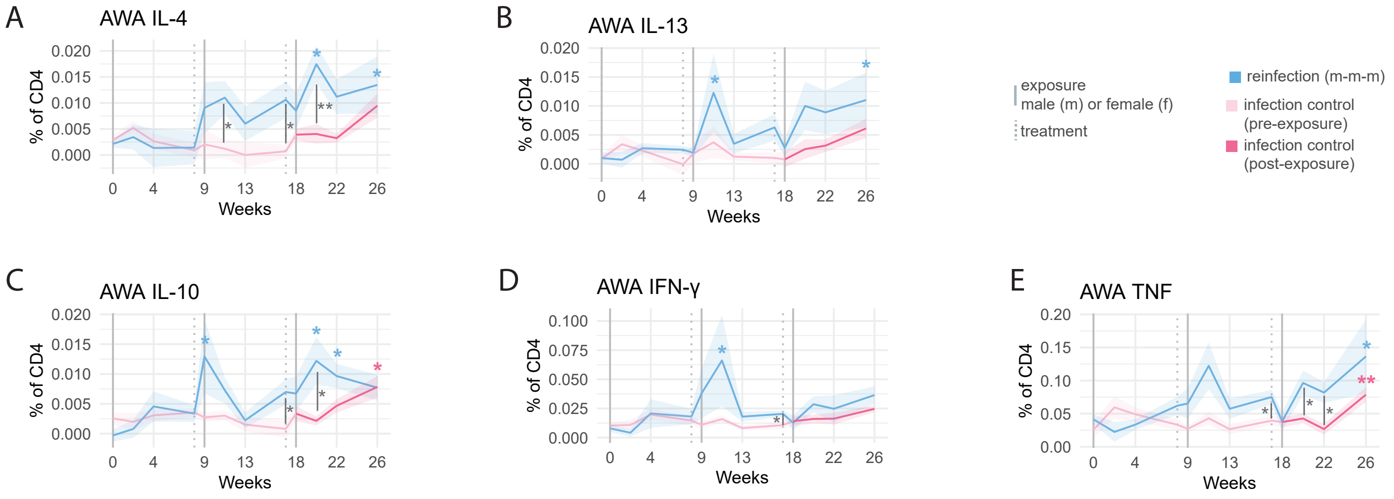


**Supp. Figure 3: Adult worm (AWA) - specific CD4 T cell responses during repeat schistosome infection.**
A-E) Ribbon plots depicting mean (lines) frequency and standard error (shaded area) of the mean of specified stimuli and populations, coloured by infection group, comparing reinfection (m-m-m) and infection control. In panels C-N a linear mixed model was performed to (separately) assess changes in the reinfection (m-m-m) and infection control group compared to the week 0 baseline, with volunteer as a random effect. FDR values are displayed in blue for reinfection (m-m-m) or red (infection control) * FDR <0.05, ** FDR<0.01. To compare between reinfection (m-m-m) and infection control groups welch's T-tests were performed at each timepoint with significant p values (uncorrected) displayed with a grey line * p <0.05, ** p <0.01. All frequencies shown are post medium subtraction.

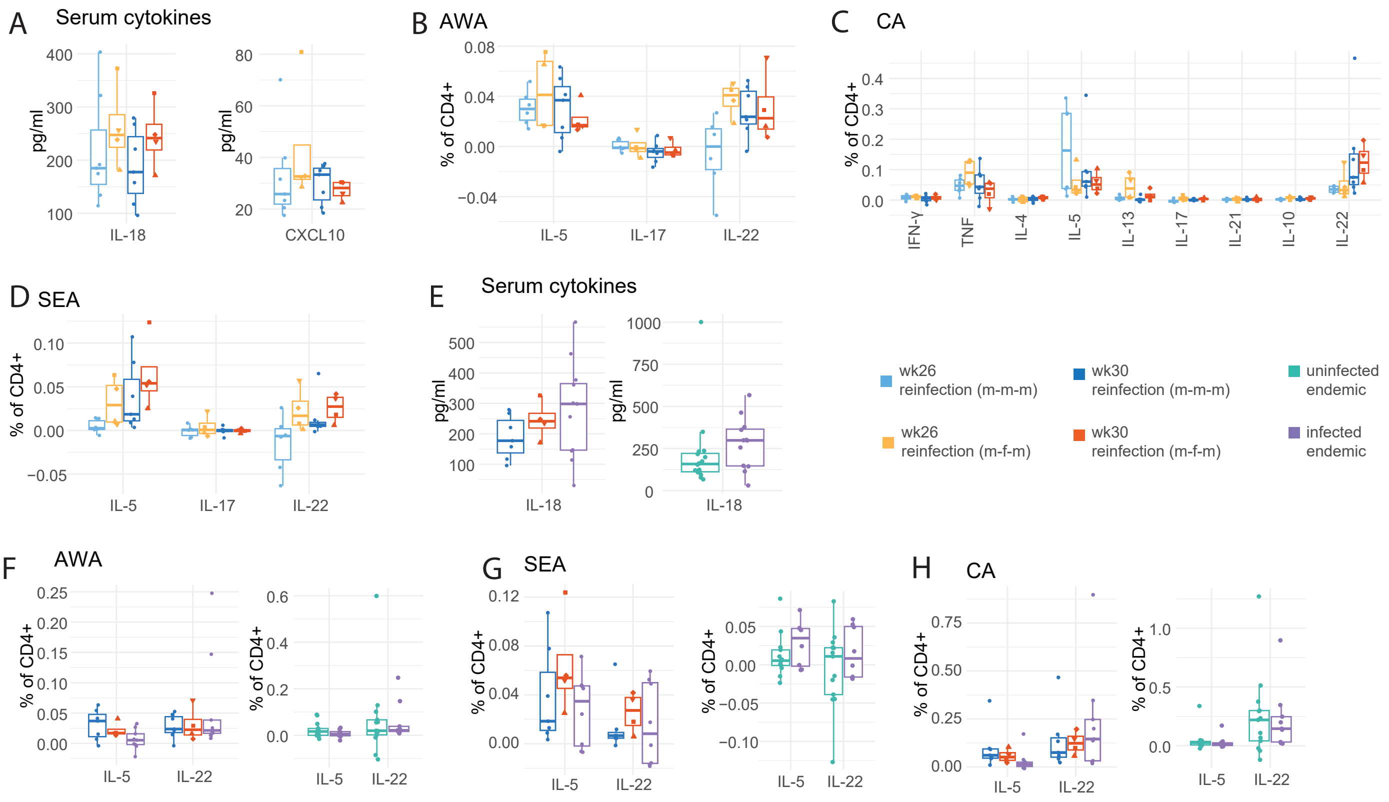


**Supp. Figure 4: Cytokine responses in controlled human infection and endemic schistosome infection.**
A) Boxplot comparing serum cytokines between reinfection (m-m-m) and reinfection (m-f-m) at week 26 and 30. C-E) Boxplots comparing AWA-specific, SEA-specific and CA-specific CD4 T cell cytokine frequency between reinfection (m-m-m) and reinfection (m-f-m) at week 26 and 30. E) Boxplot comparing serum IL-18 between controlled human infection and endemic schistosome infected and uninfected individuals. F-H) Boxplot comparing AWA-specific, SEA-specific and CA-specific CD4 T cell cytokine frequency between controlled human infection and endemic schistosome infected and uninfected individuals. All CD4 cytokine frequencies shown are post medium subtraction. Boxplots display central line (median), and hinges (25th and 75th percentile) with whiskers extending from the hinge to the largest/smallest value or 1.5x the interquartile range. No significant P values were derived from wilcoxon signed-rank tests when comparing between timepoints, or Mann-Whitney test to compare across groups.

| Prior infection | | | Current infection | | | | | |
|---|---|---|---|---|---|---|---|---|
| SEA IgG (titer) | Adult worm IgM (titer) | Combined Result (SEA IgG or AWA IgM) | CAA (pg/ml) | CAA Result | CCA Result | Kato Katz (*S. mansoni*, eggs/gram) | Kato Katz (*S. mansoni*) | Final Group |
| **128** | **512** | **Positive** | **8478** | **Positive** | **Positive** | **784** | **Positive** | **Infected** |
| <32 | **32** | **Positive** | **5998** | **Positive** | **Positive** | **624** | **Positive** | **Infected** |
| <32 | <16 | Negative | **2124** | **Positive** | **Positive** | **520** | **Positive** | **Infected** |
| **128** | **512** | **Positive** | **121** | **Positive** | **Positive** | **416** | **Positive** | **Infected** |
| **64** | **64** | **Positive** | **>10000** | **Positive** | **Positive** | **396** | **Positive** | **Infected** |
| <32 | **128** | **Positive** | **3962** | **Positive** | **Positive** | **264** | **Positive** | **Infected** |
| <32 | **32** | **Positive** | **2620** | **Positive** | **Positive** | **228** | **Positive** | **Infected** |
| **64** | **128** | **Positive** | **4776** | **Positive** | **Positive** | **208** | **Positive** | **Infected** |
| **128** | **512** | **Positive** | <OOR | Negative | **Positive** | **48** | **Positive** | **Infected** |
| <32 | **32** | **Positive** | **380** | **Positive** | **Positive** | **40** | **Positive** | **Infected** |
| <32 | <16 | Negative | **427** | **Positive** | **Positive** | **4** | **Positive** | **Infected** |
| <32 | **128** | **Positive** | <OOR | Negative | **Positive** | **4** | **Positive** | **Infected** |
| <32 | <16 | Negative | <OOR | Negative | Negative | 0 | Negative | Uninfected |
| <32 | <16 | Negative | <OOR | Negative | Negative | 0 | Negative | Uninfected |
| <32 | <16 | Negative | <OOR | Negative | Negative | 0 | Negative | Uninfected |
| <32 | <16 | Negative | <OOR | Negative | Negative | 0 | Negative | Uninfected |
| <32 | <16 | Negative | **16** | **Positive** | Negative | 0 | Negative | Uninfected |
| <32 | <16 | Negative | <OOR | Negative | Negative | 0 | Negative | Uninfected |
| **64** | **512** | **Positive** | <OOR | Negative | Negative | 0 | Negative | Uninfected |
| <32 | <16 | Negative | <OOR | Negative | Negative | 0 | Negative | Uninfected |
| <32 | **128** | **Positive** | <OOR | Negative | Negative | 0 | Negative | Uninfected |
| <32 | **32** | **Positive** | <OOR | Negative | Negative | 0 | Negative | Uninfected |
| <32 | <16 | Negative | <OOR | Negative | Negative | 0 | Negative | Uninfected |
| <32 | **512** | **Positive** | <OOR | Negative | Negative | 0 | Negative | Uninfected |
| <32 | <16 | Negative | <OOR | Negative | **Positive** | 0 | Negative | Uninfected |
| <32 | <16 | Negative | <OOR | Negative | Negative | 0 | Negative | Uninfected |
| <32 | <16 | Negative | <OOR | Negative | **Positive** | 0 | Negative | Uninfected |
| <32 | <16 | Negative | <OOR | Negative | Negative | 0 | Negative | Uninfected |

**Supp. Figure 5: Prior and current infection status in endemic individuals.**
Multiple diagnostic methods were used to establish prior and current schistosome infection status in endemic individuals. To establish prior infection anti-SEA IgG reactivity (cutoff <32) and anti-adult worm IgM (cutoff <16) titers were measured. Current infection status was determined using a combination of the serum UCP-LF CAA assay (CAA, cutoff 10 pg/ml), the rapid diagnostic test CCA assay (in urine) and Kato Katz to find schistosome eggs in stool. Volunteers were designated into the infected group when 2/3 current infection diagonistics (CCA, CAA and Kato Katz) were positive.

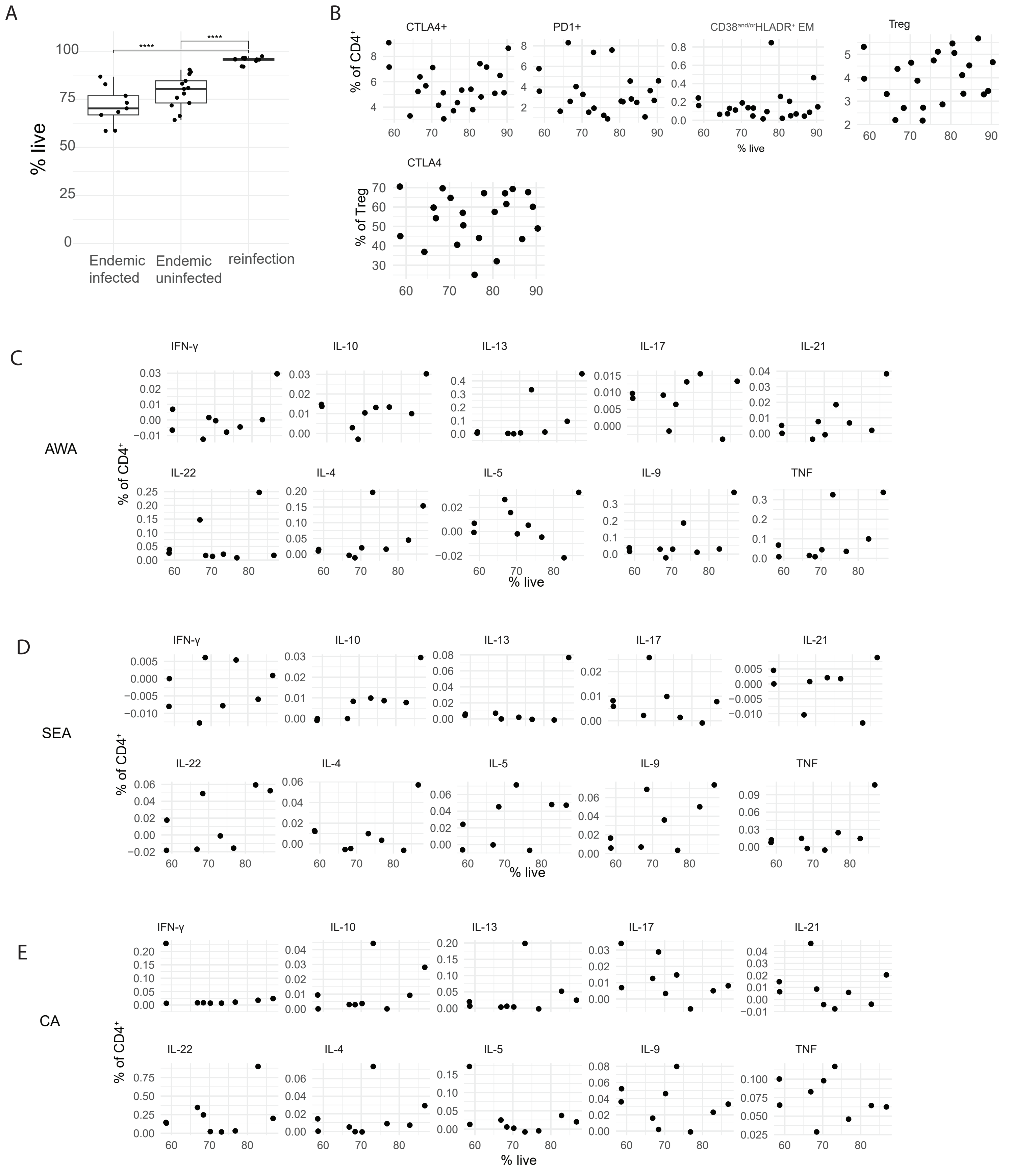


**Supp. Figure 6: Viability**

Viability, determined during flow cytometric staining via the use of an amine reactive dye. A) Average viability between 4 conditions (media, AWA, SEA, CA) is shown, split by participant grouping. Differences between groups were determined via ANOVA with tukey's HSD testing. **** p<0001. B) Scatter plot comparing unstimulated cell population frequency and viability. C-E) Scatter plots comparing AWA stimulated (A), SEA-stimulated (B) and CA-stimulated (C) cytokine frequency (as % of $CD4^+$ T cells) to viability. Correlation between viability and cell population frequency was performed using Spearmans correlation with resultant p values adjusted for multiple corrections using the FDR method. No significant correlations were found (FDR>0.05).

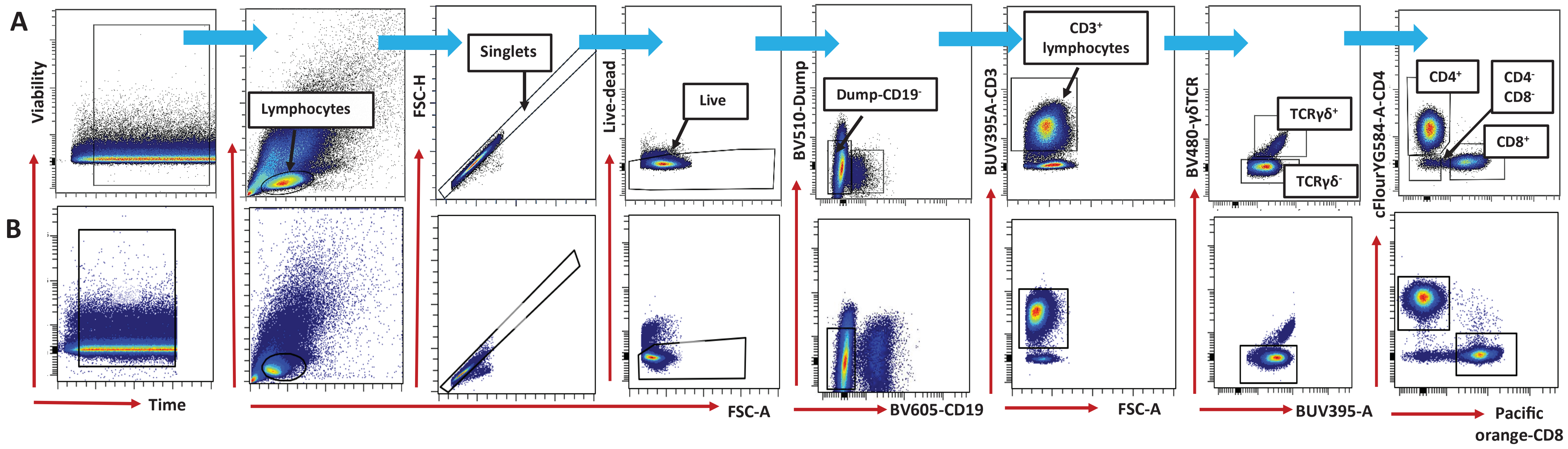


**Supp. Figure 7. Gating scheme to define CD4$^{+}$ and CD8$^{+}$ T cell populations.** PBMC samples assessed by flow cytometry following stimulation or unstimulated and sequentially gated as show above to define viable cells, lymphocytes, singlets, live, CD3$^{+}$, TCRγδ$^{-}$ lymphocytes and CD4$^{+}$ T cells in the reinfection (**A**) and endemic (**B**) PBMC samples.

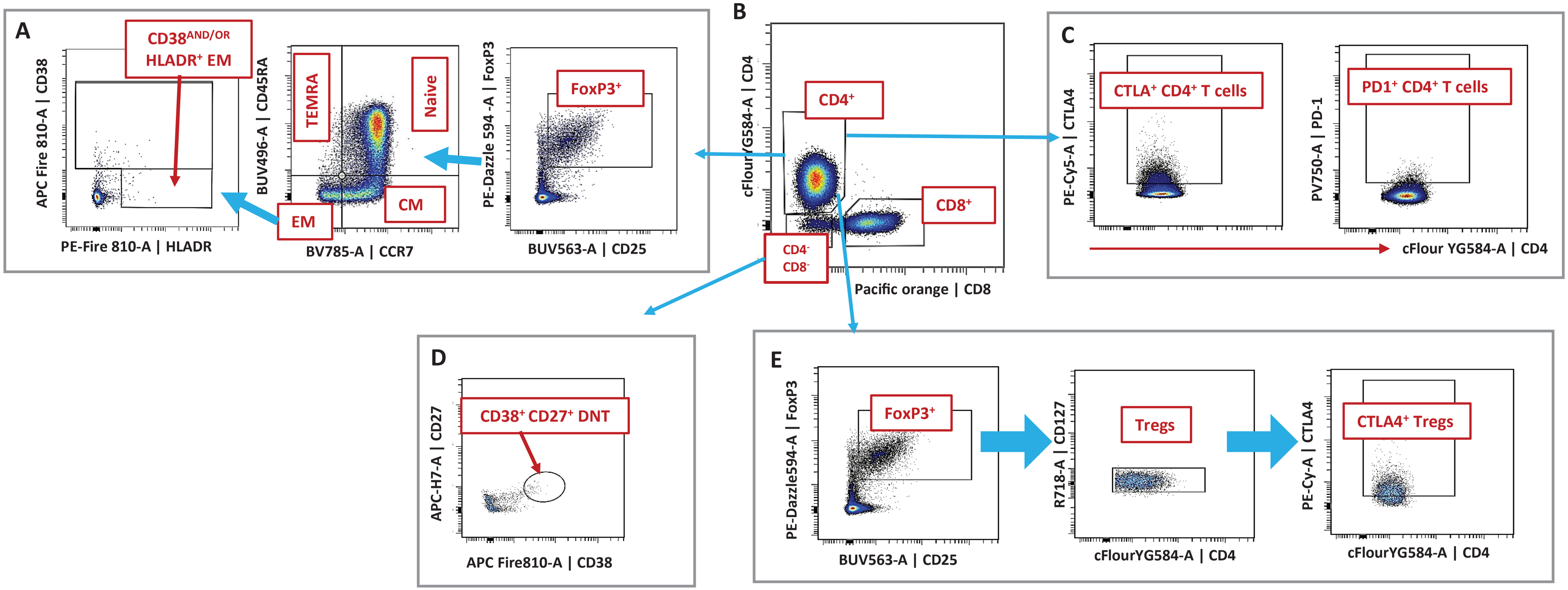


**Supp Figure 8. Gating scheme used to define cellular response in unstimulated PBMC samples from reinfection (m-m-m and m-f-m) and endemic volunteers.** Gating for; **A.** CD38$^{AND/OR}$HLDR+ T cells from EM subset, **B**. CD4$^-$ CD8$^-$ (DN), CD4$^+$ and CD8$^+$ T cell subsets, **C.** gating for CD4$^+$ T cell phenotypes CTLA$^+$ and PD1$^+$ CD4$^+$ T cells **C.** gating to define CTLA+ T regs and **D**. CD38$^+$ CD27$^+$ double negative T cells from CD4- CD8- population, and **E**. CTLA4$^+$ expressing regulatory T cell gate from Treg and FoxP3$^+$ subsets. **EM;** effector memory, **DN;** Double negative, **CM;** central memory, **TEMRA;** terminally differentiated effector memory cells

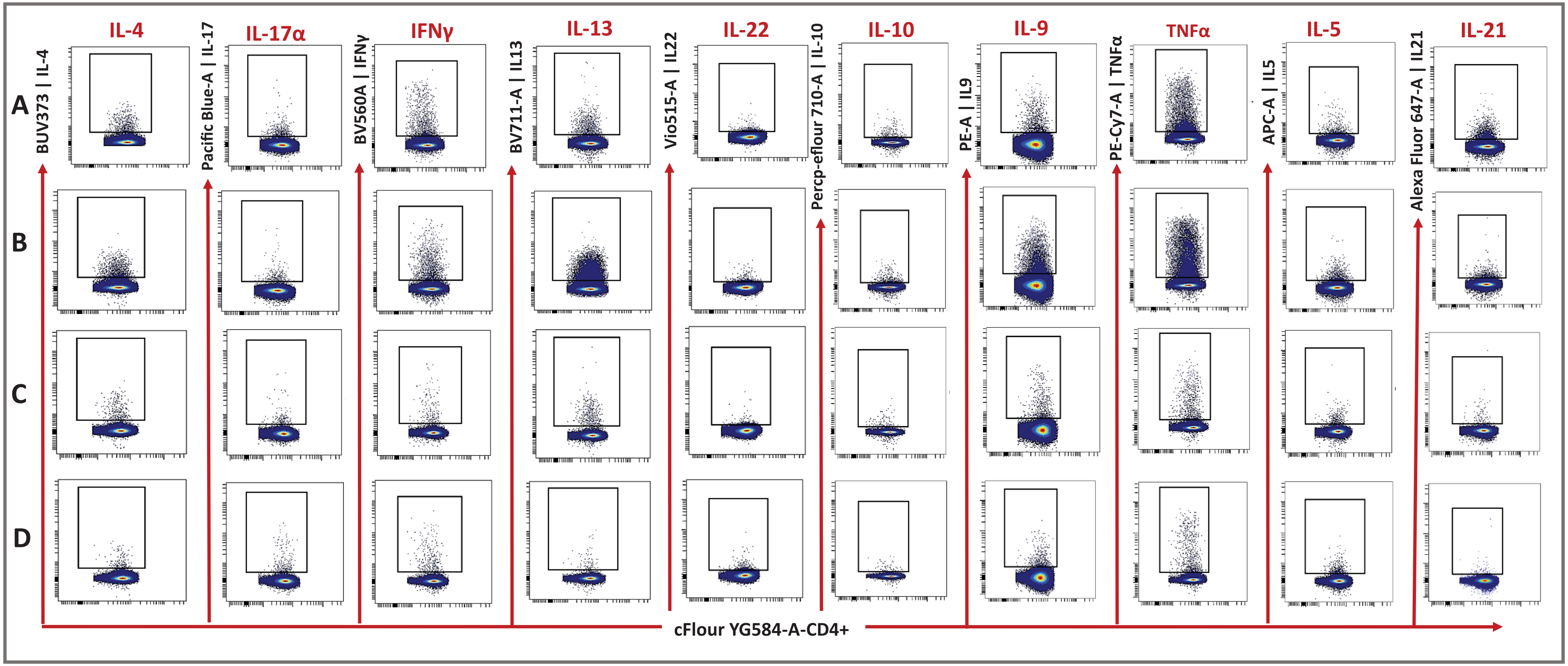


**Supp. Figure 9. Representative gates for CD4⁺ T cell-specific cytokine response post-stimulation in the four study sub-groups.** CD4⁺ T cell pre-gating for cytokine response. **A-D**. gating scheme for cytokine response in the m-m-m reinfection group (**A**), m-f-m reinfection group (**B**), endemic infected (**C**) and endemic uninfected (**D**) groups. Each gate represent cytokine gate concatenated for AWA, CA, SEA and MED stimulated samples
